# Defining use cases for biomarkers and tests across tuberculosis infection, disease and treatment: An international consensus and prioritisation exercise

**DOI:** 10.64898/2026.09.01.26361598

**Authors:** Sacha Wright, Federico Fama, Angus de Wilton, Ellen Steward, Francesca Saluzzo, Chiara Sepulcri, James Greenan-Barrett, Khay Mar Aung, Thi Mai Nguyen, Nora Engel, Seda Yerlikaya, Hanif Esmail, Salome Charalambous, Cecily Miller, Ruvandhi Nathavitharana, Luan Nguyen Quang Vo, Mikashmi Kohli, Delia Goletti, Daniela Cirillo, Mahdad Noursadeghi, Claudia Maria Denkinger, Emily Lai-Ho MacLean, Rishi K Gupta, Ankur Gupta-Wright

## Abstract

**Background:** Translation of tuberculosis (TB) biomarker and diagnostic research into tools that improve patient and public health outcomes has been slow, partly because no internationally agreed framework exists defining the use cases that new biomarkers and tests should address. We aimed to identify, validate, and prioritise use cases for TB biomarkers and tests across *Mycobacterium tuberculosis* (*Mtb*) infection, TB disease, TB treatment, and post-TB care, through an international consensus process.

**Methods and Findings:** We conducted a scoping review of the literature, guidelines, and target product profiles (24 documents; 69 candidate use cases consolidated to 13), followed by a hybrid RAND/UCLA modified Delphi consensus process involving 185 identified interest-holders, including clinicians, researchers, diagnostic developers, industry, funders, civil society, national TB programmes, and policymakers (including WHO representatives). Interest-holders completed an online survey rating agreement with each use case, and attended a consensus meeting to discuss use cases with less than 80% agreement, followed by a further validation meeting. Eleven use cases were retained across four TB care pathway stages: three for *Mtb* infection, four for disease detection, three for treatment optimisation, and one for post-TB care. Highest-priority use cases were detection of drug-resistant TB, prediction of progression from infection to disease, identification of current *Mtb* infection, and improved diagnosis of active TB disease.

**Conclusions:** This consensus exercise provides the first comprehensive, prioritised framework of use cases for TB biomarkers and tests, spanning the full care pathway. These eleven priority use cases can guide investment, focus biomarker discovery, and inform future target product profiles, funding calls, and policy development. Applying the framework to the current biomarker pipeline is a key next step to address gaps between innovation and priority needs.

## Background

Despite improved diagnostics and effective treatment, tuberculosis (TB) remains a leading cause of global morbidity and mortality, with an estimated 10.7 million incident cases and 1.1 million deaths due to TB in 2024.^[1]^ Progress towards achieving the End TB Strategy targets has been slow,^[2]^ and approximately one quarter of people with TB disease remain undiagnosed and therefore untreated.^[1]^ Closing diagnostic and treatment gaps is essential to reducing disease transmission, preventing avoidable deaths, and achieving TB elimination goals.

Important unmet needs persist across the full spectrum of *Mtb* infection and TB disease. Around one quarter of the world’s population is estimated to have an immune response to *Mycobacterium tuberculosis (Mtb)* ^[3]^, yet current tools cannot reliably identify the small proportion of infected individuals at risk of progression to disease, limiting the efficiency of preventive strategies.^[4]^ Among those diagnosed with TB disease, treatment is largely standardised and prolonged, despite increasing evidence that many individuals could be cured with shorter regimens.^[5,6]^ Therefore, better tests for defining TB states, assessing infectiousness, predicting progression, guiding treatment, and monitoring response could meaningfully improve both individual outcomes and wellbeing, and programme-level impact.

Over the past decade, TB biomarker discovery and diagnostic research have expanded considerably.^[7,8]^ However, translation into tools that are implemented at scale in TB programmes has been limited. Many promising biomarkers and tests fail to progress beyond early development because their comparative diagnostic performance, cost-effectiveness or implementation feasibility is suboptimal compared to available options; however, even potentially superior biomarkers and tests fail to progress because their intended clinical or programmatic role is unclear, they are poorly aligned with health system needs or health financing capacities, or they remain insufficiently prioritised by end users and policymakers.^[9,10]^ This lack of alignment contributes to inefficiencies in research and development, and limits the population-level impact of innovation.^[11]^

Target product profiles (TPPs) have helped articulate desired characteristics for new TB diagnostics and align product development with public health priorities, and TB benefits more from an established TPP landscape than many other disease areas.^[12]^ Nonetheless, potential use cases for biomarkers remain and fragmented and inconsistently defined across different TPPs, guidelines, and fast-evolving academic literature. Additionally, there has not been a systematic effort since 2014 to assess which use cases should be prioritised for investment to maximise impact with engagement of broad interest-holder groups.^[13,14]^

## Methods

We sought to address this gap in the TB Biomarker and Test Landscape (TB-BIOATLAS) project by providing a consolidated, consensus-driven, and prioritised framework for use cases for biomarkers and tests for TB. We aimed to identify and map potential use cases for TB biomarkers across TB stages (*Mtb* infection, TB disease, TB treatment, and post-TB care); to develop consensus among interest-holders regarding the validity of these use cases; and to prioritise use cases to inform future biomarker and diagnostic research, funding, and product development.

We used a multi-step, iterative process to identify, validate, and prioritise use cases for TB biomarkers **(Fig 1)** (protocol published via Wellcome Open ^[15]^). Firstly, we undertook a scoping review to identify potential use cases already described in the literature, including guidelines and TPPs. Secondly, we undertook a hybrid RAND/UCLA appropriateness method/modified Delphi consensus process with diverse interest-holder groups to establish agreement and validate putative use cases, including a survey and consensus meeting.^[16,17]^ Thirdly, we asked interest-holders to prioritise use cases. Methodology adhered to ACCORD guidelines where appropriate, and scoping review reporting adhered to PRISMA-ScR.^[18]^ Our methods have been previously described in more detail.^[15]^

**Fig 1:**
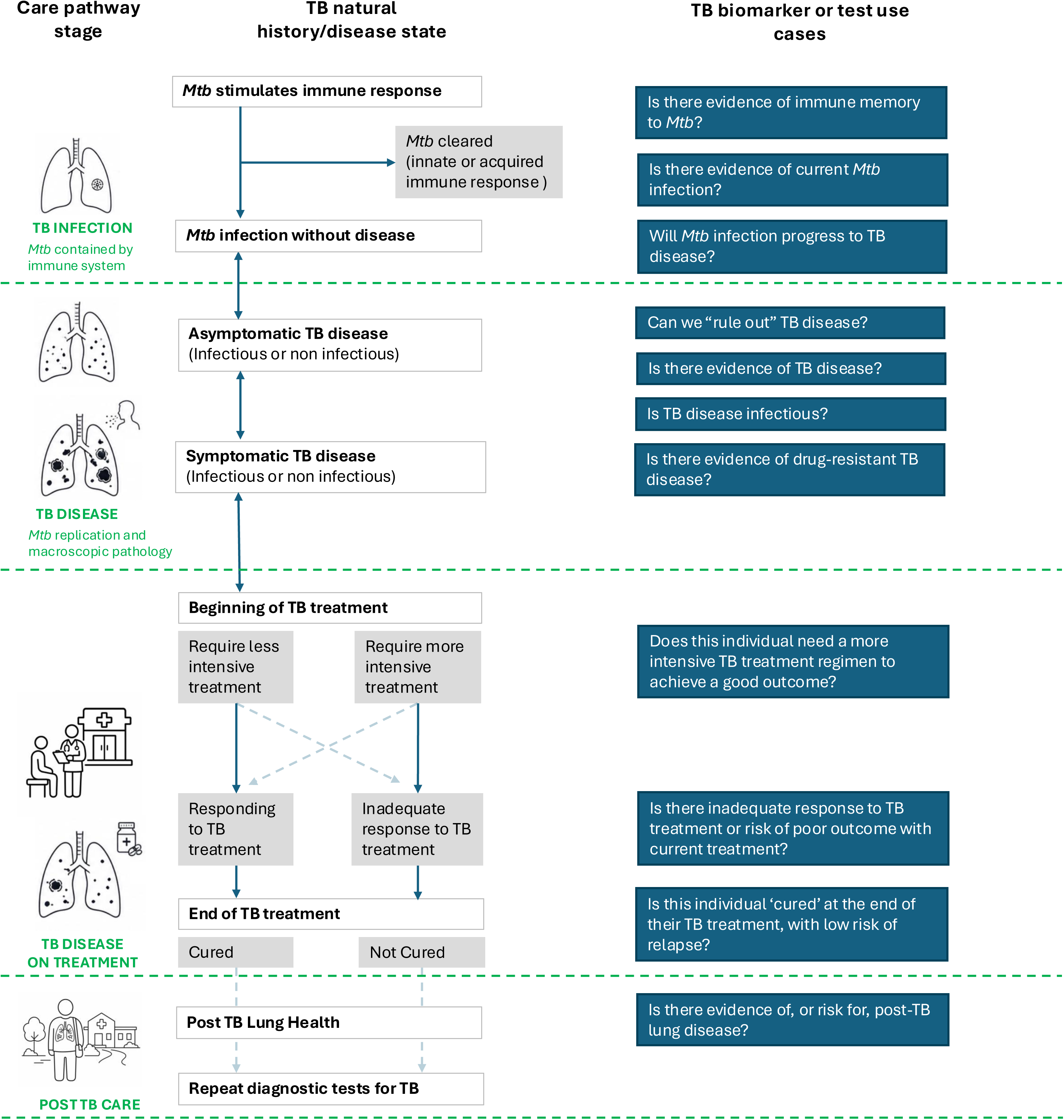
Overview of the adapted Delphi consensus process and study participants. Biomarkers can be defined as ‘characteristics that can be measured as an indicator of a biological process, pathological process or response to an exposure or intervention’.^[8]^ They are broad and can be derived from biochemical, molecular, genetic, pathological, radiographic, clinical or physiological measurements or characteristics, and their measurement should be objective and repeatable. They can also be further defined according to their intended use (e.g. diagnostic biomarker, predictive biomarker, monitoring biomarker). In the context of biomarkers or tests, a use case can be defined as a ‘specific scenario or instance where a biomarker or test provides actionable information that can improve patient care or public health outcomes’.^[23]^

### Scoping review

We searched PubMed/MEDLINE, Global Index Medicus, WHO IRIS, and the WHO and FIND websites on July 21, 2025, to identify descriptions of TB biomarker use cases (we defined use cases as scenarios where a biomarker or test provides actionable information that can influence management and/or impact outcomes). We were particularly interested in identifying TPPs and international guidelines where use cases were likely to be described. Our search included terms for ‘tuberculosis’, ‘TPP’, and ‘guidelines’ (full list of search terms and databases can be found in **S1 Table**; inclusion and exclusion criteria can be found in **S2 Table** and **S3 Table**).^[15]^ There were no time limits on the search, but where multiple versions of the same document were available, only the most recent was included for final data extraction. After de-duplication, citations were screened using Rayyan software by two independent reviewers at title/abstract and full-text stages. Conflicts were resolved through discussion between reviewers, with arbitration by a third reviewer where required. Reasons for exclusion were recorded at the full text review stage.

Data from included sources were extracted using a custom-designed extraction form in an Excel spreadsheet and synthesised descriptively into use cases (**S4 Table**). Where available, we extracted example tests for a particular use case, as well as potential impact on patient- and population-level health outcomes for that use case. Similar or overlapping use cases identified from multiple sources were consolidated into a list of distinct use cases or were considered example ‘sub-use cases’ (e.g. if they described application in a specific setting or population group). The list of main use cases was then mapped to TB stages and steps in the TB care pathway, which were adapted from the WHO documents and the ICE-TB framework.^[19–21]^

### Participants/Interest-Holders

The project leads (RKG, ELM, AGW, SW) managed and conducted the consensus process. Categories of interest-holder groups included clinicians, TB programme staff, policymakers and NGOs, researchers (including discovery science, epidemiology and clinical research), industry and test developers, and TB community and patients. They were identified through existing professional networks, publicly accessible online sources, and snowball recruitment **(S5 Table)**.^[15]^

The project team reviewed the list of potential interest-holders to ensure diversity and recentness of contact details and on this basis selected a subset to invite as participants. Invitations were sent to potential interest-holders until at least eight to ten individuals were identified for each group. For interest-holders accepting participation, information on conflicts of interest, demographics, relevant experience, and geographic regions was recorded. Additional interest-holders were invited when necessary to ensure a balance of gender and geographic region across groups. Interest-holders were given 6 weeks to respond to the survey. Alongside completing the survey, they were invited to attend a hybrid in-person/online consensus meeting in November 2025.

### Consensus process

Following the scoping review that identified candidate use cases, the consensus process included an interest-holder survey and two facilitated consensus meetings (**see Fig 1**). The use cases extracted from the scoping review informed the questions for an online survey, which were distributed to participating interest-holders by email after piloting and feedback within the project team (**S6 Appendix)** ^[15]^. The survey presented the proposed use cases organised into four stages of TB (*Mtb* infection, TB disease, TB treatment and post-TB), and asked respondents to anonymously indicate agreement with each use case using a five-point Likert scale.^[15]^

Survey responses were collected between Oct 28, 2025, and Dec 19, 2025. Respondents who indicated they did not agree with a particular use case were asked to provide free-text comments to explain why. In survey analysis, responses of ‘agree’ or ‘strongly agree’ were considered as agreement. The consensus threshold was set at 70%, with use cases reaching <80% discussed further and >80% included, as per established consensus thresholds.^[16]^ Free-text comments were analysed qualitatively to identify common themes and concepts, and Likert scale-style questions were analysed and ranked by interest-holder group using R Studio 4.4.2.^[22]^ To capture variability of agreement across diverse interest-holders, survey response analysis was also stratified by group.

The findings of the scoping review, survey results, and key themes from comments were presented at the interest-holder hybrid consensus meeting. Use cases with <80% agreement were discussed in an open forum. At the end of each discussion, participants voted on whether the use case was valid and should be included in the final framework using anonymous online polling software. The project team did not participate in voting. The list of use cases was then further adapted, and/or language of the use cases fine-tuned based on interest-holder input and polling results. It was decided *a priori* that a further round of surveys would only be required if consensus was not achieved through the initial survey and meeting. The final use cases, including an overview of any amendments to the framework, were presented at a final online consensus meeting in March 2026.

### Prioritisation of use cases

To understand which use cases are considered high priority for development, we asked interest-holders to prioritise use cases using an online survey.^[15]^ After indicating agreement with use cases, interest-holders were presented with the use cases again and asked to rate the importance of each as ‘high’, ‘medium’ or ‘low’, with a maximum of 5 use cases permitted to be rated as ‘high importance’. The proportion of respondents indicating a use case was of ‘high’ importance was calculated per interest-holder group, and an overall ranking score was created based on the median percentage of use cases ranked as “high importance” across groups. Results of the prioritisation of use cases were presented at both interest-holder meetings for feedback and validation.

### Ethical considerations

This study involved a scoping review, anonymous survey, and structured consensus process with interest-holders. In line with published guidance on consensus methods, formal ethical approval was not required as participants contributed in a professional capacity; no identifiable, sensitive personal data were collected and responses were anonymized.

## Results

### Scoping review

The scoping review identified 24 documents describing use cases for TB biomarkers or tests, with most documents being WHO guidelines (n=8), TTPs (n=5), or WHO reports (n=5) (**S1 Figure, S7 Table**). 69 use cases were extracted, but with considerable inconsistency and overlap in how they were described. After consolidating overlapping use cases, 13 distinct use cases were described. These main use cases were reviewed to refine phrasing and clarity and then mapped to TB states and steps in the care pathway: three use cases related to *Mtb* infection, five related to detection of TB disease, three related to treatment of TB disease, and two related to post-TB care (**S8 Table**). Additionally, eleven sub-use cases were identified, which were applicable only to specific settings or populations described within a main use case. For brevity, use cases will herein be referred to by their shortened titles, which are detailed in **S8 Table.**

### Consensus process

From the list of 185 interest-holders identified, 166 (90%) were invited as participants, of whom 83 (45%) expressed initial interest. 88 (70 original respondents, 18 additional interest-holders invited to balance demographic characteristics) responded to the survey; participation was 43% (88/203). Survey respondents were well balanced across key characteristics including gender, geographic region, country income level, and TB burden (**S9 Table)**. The highest response rate was from researchers, but all interest-holder groups had at least twelve respondents. The survey established clear consensus (>80% agreement) for 9/13 use cases, with two use cases falling below the 70% consensus threshold, and two use cases achieving between 70-80% agreement, with significant feedback in comments (**S2 Figure**). Survey written comments are summarised in the appendix (**S10 Table**).

43 (23%) of the original interest-holders attended a hybrid in-person/online consensus meeting in November 2025, alongside an additional 5 interest-holders identified through expert suggestion. At the meeting, results of the scoping review and survey were presented to participants, with discussion of all use cases and detailed discussions for the four use cases with <80% agreement in the survey (‘exposure to *Mtb*’, ‘asymptomatic treatment’, ‘infectiousness’ and ‘relapse or re-infection’). Participation in polling ranged from 71% to 79% of attendees.

Based on anonymous online polling and discussion, two use cases were excluded from the final framework (‘asymptomatic treatment’ and ‘relapse or re-infection’), one use case was retained but revised for clarity (‘exposure to *Mtb*’), and one-use case was retained as originally described (‘infectiousness’). Key messages from interest-holder discussions are detailed in **S10 Table.** The final set of use cases was agreed in the second interest-holder meeting.

### Use cases for TB biomarkers and tests by TB care pathway stage

Based on the scoping review and multi-step, iterative consensus process, we identified 11 use cases for biomarkers across the four TB care pathway stages (**Fig 2**). Descriptions of use cases and their potential patient- and population-level health outcomes can be found in **Table 1**. Interest-holders’ survey comments and discussion at the consensus meeting highlighted that tests could also be useful for research outcomes, which could in turn have potentially important public health impact (i.e., ‘current *Mtb* infection’ providing useful information as potential surrogate endpoint for ‘prevention of *Mtb* infection’ vaccine trials). These potential research outcomes are also described in **Table 1**.

**Fig 2:**
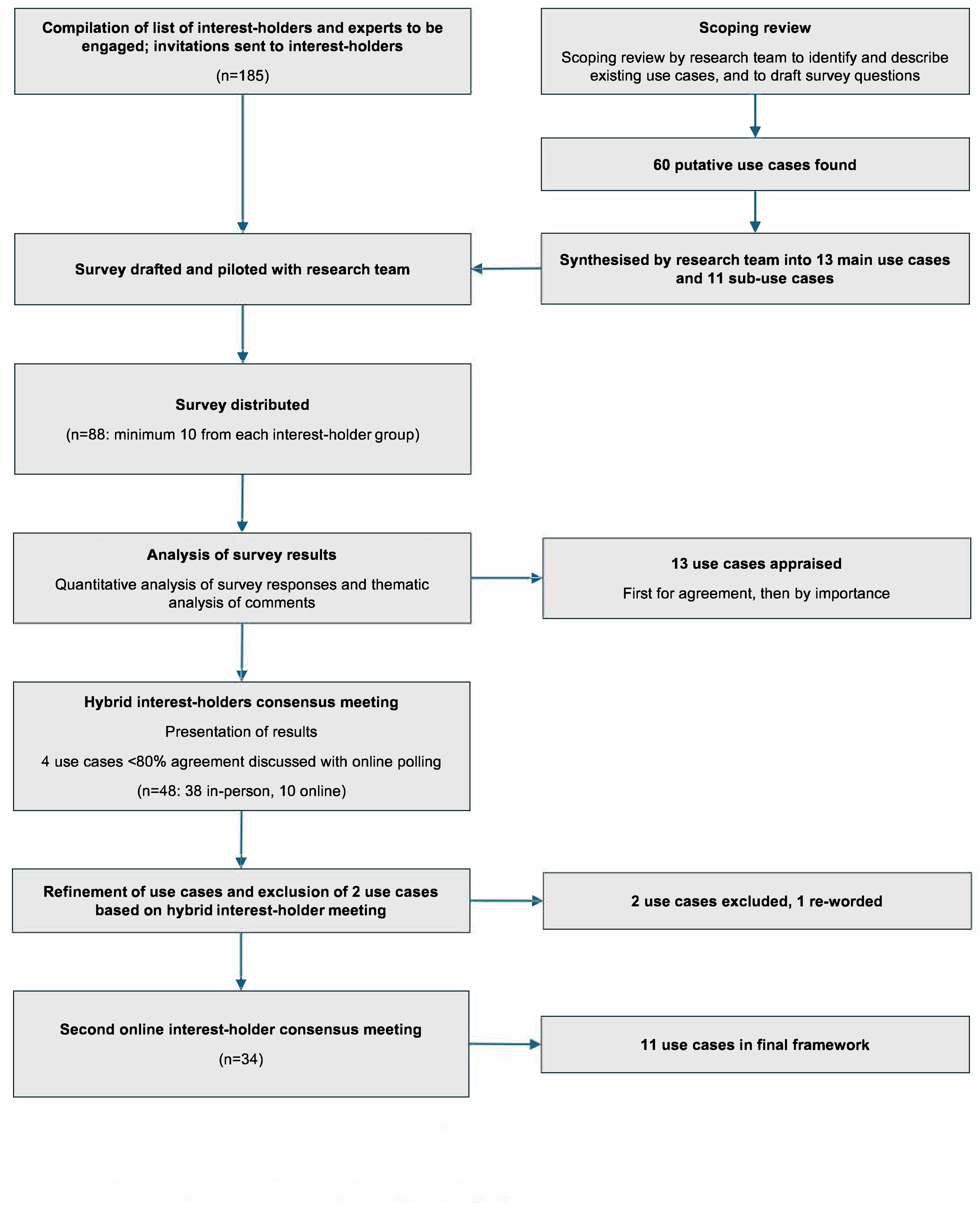
Use cases for tuberculosis biomarkers and tests. This figure depicts the biomarker and test use cases identified through the consensus process, mapped across the major stages of the tuberculosis (TB) care pathway (*Mtb* infection, TB disease, TB disease on treatment, and post-TB care) and the corresponding natural history and disease states of TB. The framework spans the continuum from Mycobacterium tuberculosis (Mtb) infection without disease, through asymptomatic and symptomatic TB disease, diagnosis and treatment, treatment response and outcomes, and post-TB lung health and disease. Use cases are positioned alongside the disease states, transitions, or clinical decision points that they are intended to detect, predict, monitor, or influence. Disease states and terminology were aligned, where possible, with definitions from the International Consensus for Early-Stage Tuberculosis (ICE-TB) and the World Health Organization (WHO). Solid arrows indicate the usual route of transition between states, while dashed arrows indicate possible (alternative) routes. Progression between TB states is dynamic and not necessarily unidirectional.

**Table 1:**
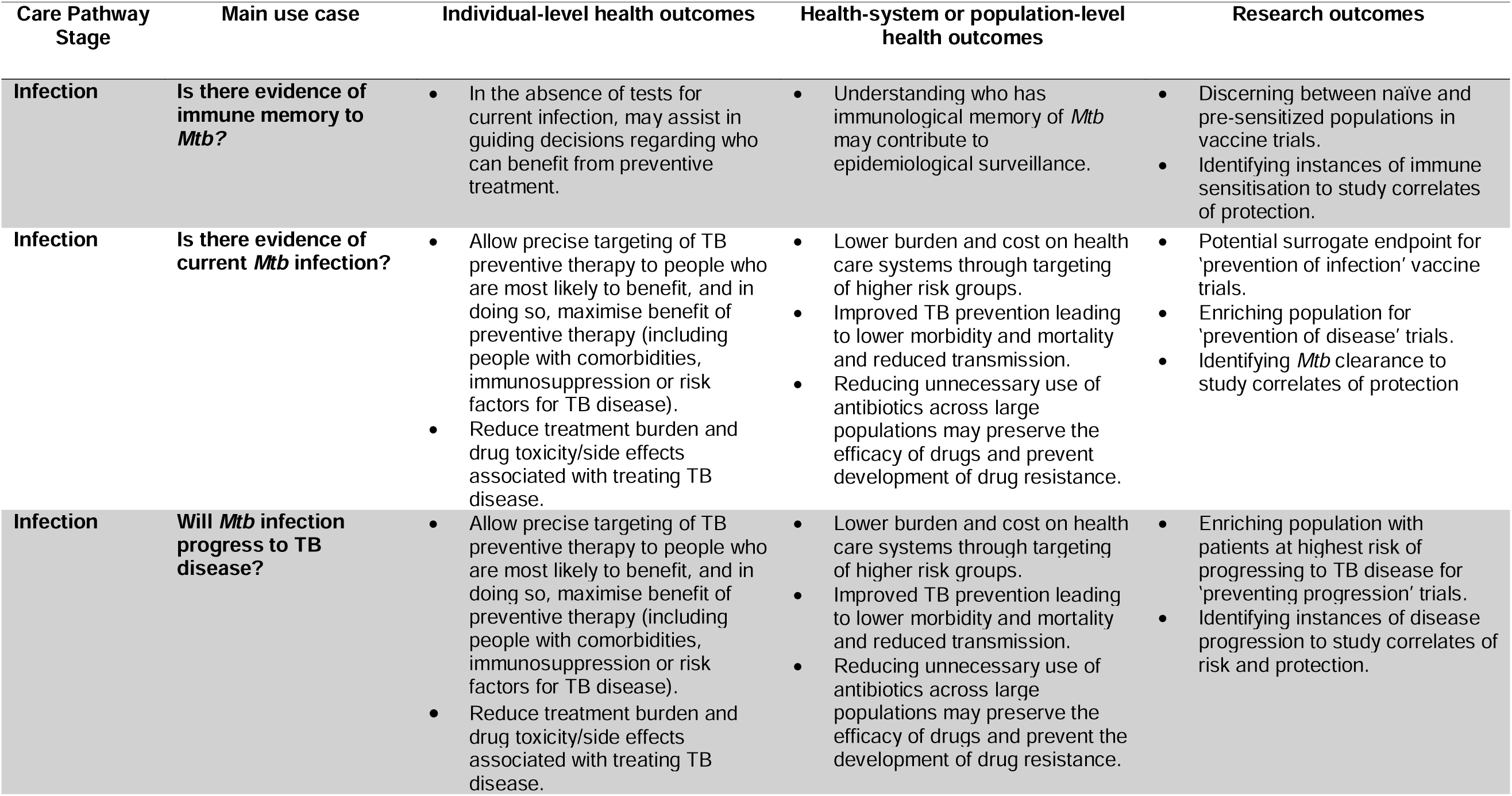
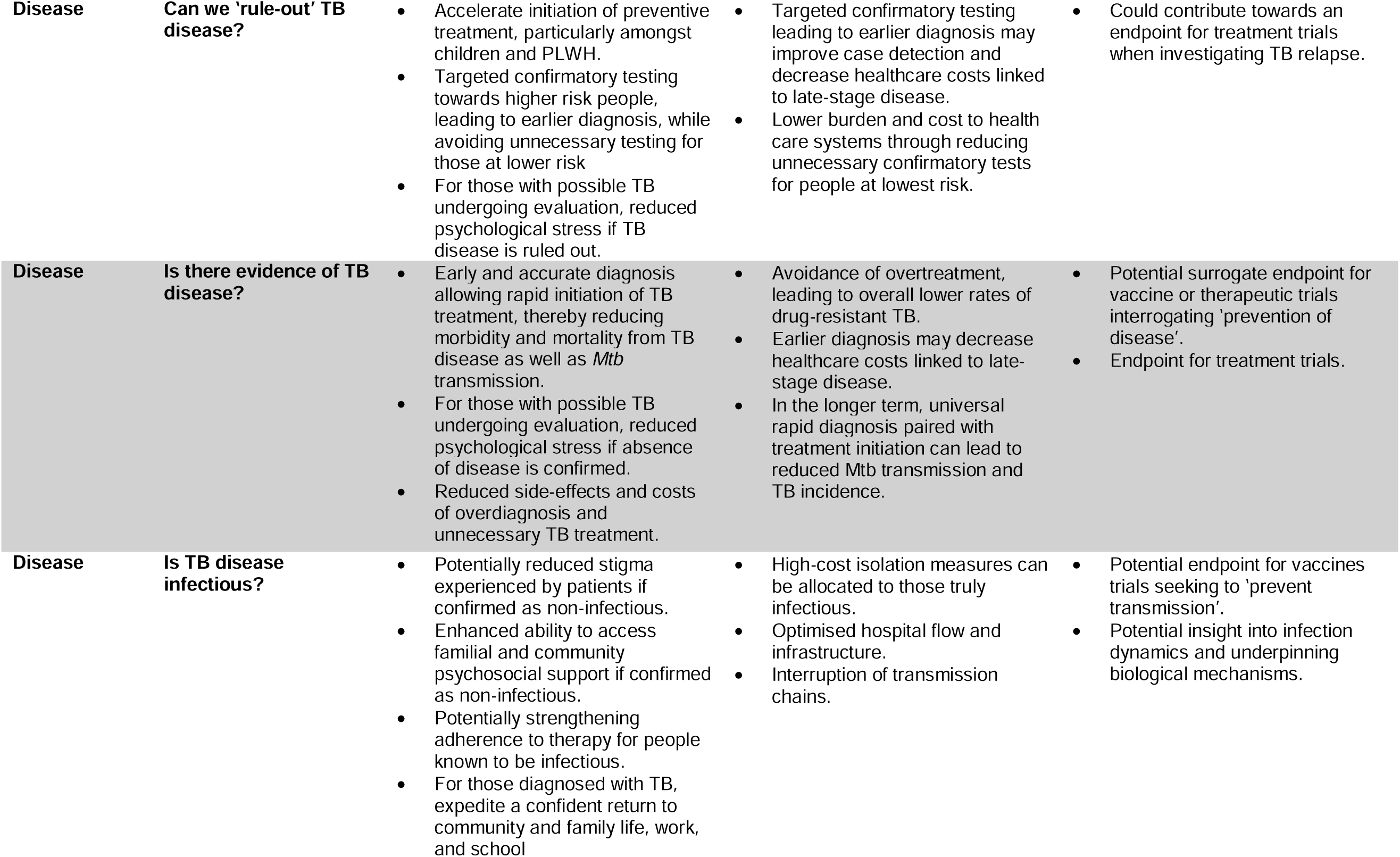
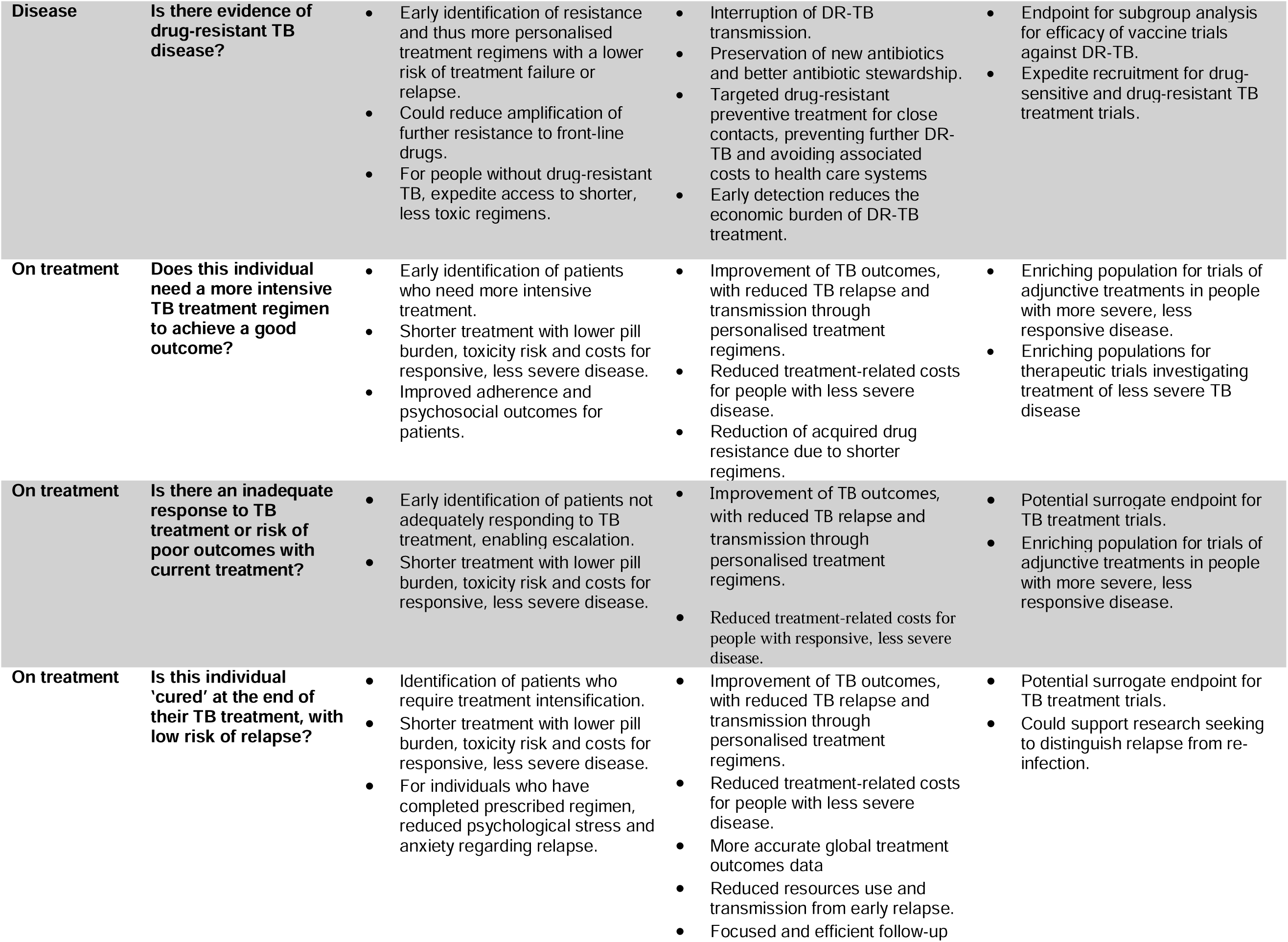
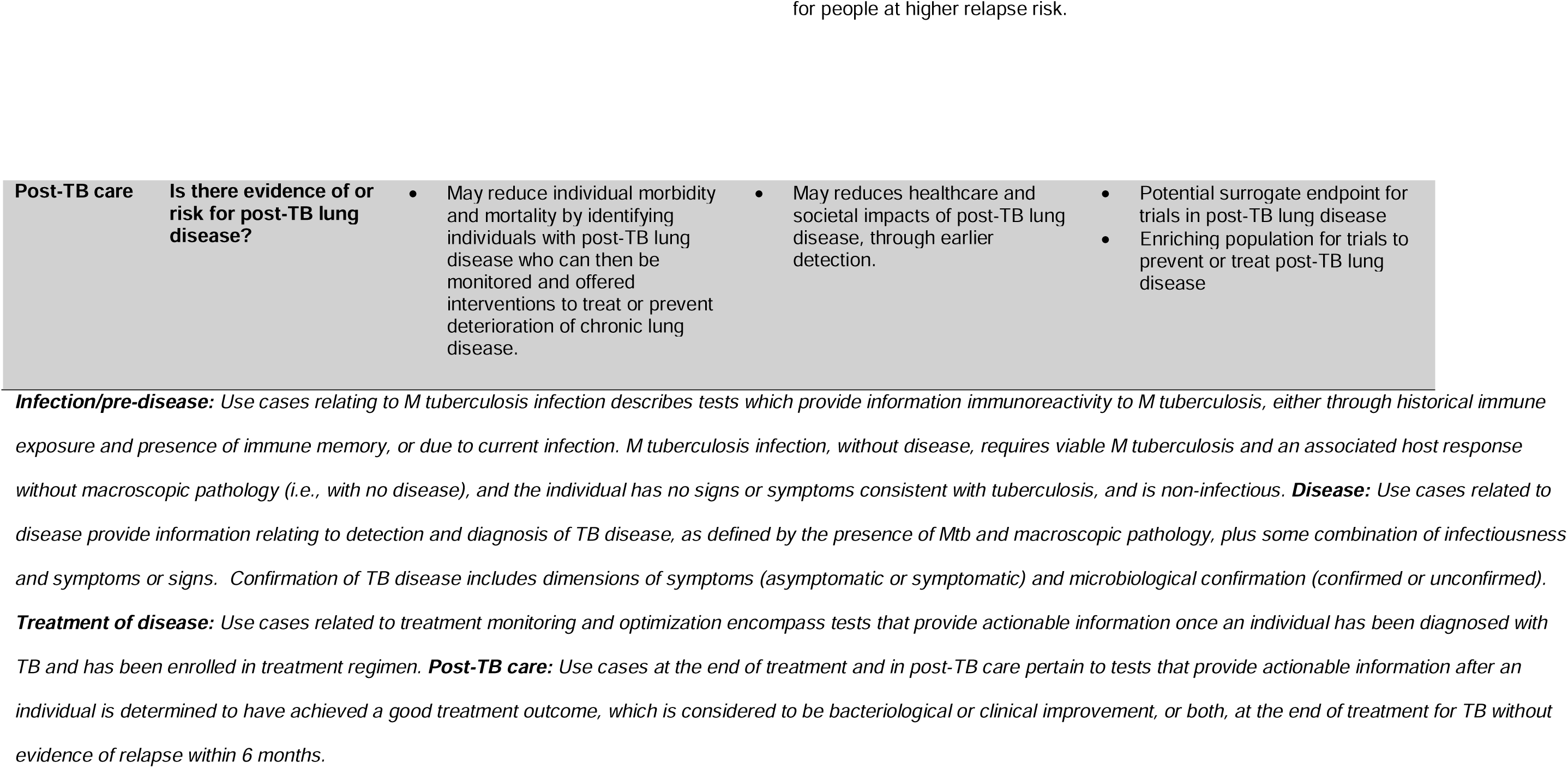
Main use cases, with associated potential patient- and population-level health outcomes and research outcomes.

#### Mtb infection stage

Conceptually, the *Mtb* infection stage describes the time during which there is presence of viable *Mtb* in the host in the absence of pathology, signs or symptoms associated with TB disease.^[20]^ Three use cases were identified and validated for the infection stage (**Fig 2).**

##### Is there evidence of immune memory to Mtb?

Currently, immunosensitisation assays such as interferon-γ release assays (IGRA) or Tuberculin Skin Tests (TST) are widely used as proxies for *Mtb* infection.^[24]^ These tests may reflect host immunosensitisation to *Mtb*, but cannot differentiate current from historical infection that has been cleared through host immune responses or treatment,^[25]^ and are not adequately predictive of progression to TB disease or benefit from TB preventive treatment (TPT).^[24]^ Immunosensitisation was identified as a potential use case from the scoping review (‘exposure to *Mtb’*). However, consensus for this use case was not reached, with agreement on the survey being 67%, and interest-holders at the consensus meeting emphasising a higher-priority need to identify current, viable *Mtb* infection to better target interventions such as TPT, or to support TB vaccine trials by providing a measurable endpoint. At the consensus meeting, participants voted to exclude the use case from the final framework (23/38 [61%]). Nevertheless, interest-holders also supported the distinction of viable *Mtb* infection from immunoreactivity. Hence, to avoid conflation between tests for immunoreactivity and those for viable infection while recognising the need to move beyond the status quo, the *‘*exposure’ use case was retained and revised as *‘Is there evidence of immune memory to Mtb’,* alongside the distinct ‘current *Mtb* infection’ use case.

##### Is there evidence of current Mtb infection?

‘Evidence of current *Mtb* infection’ was identified as a use case for a biomarker, with high agreement in surveys (89%) and interest-holder meetings, although the absence of a gold standard for viable *Mtb* infection was identified as a key challenge.

##### Will Mtb infection progress to TB disease?

A biomarker to predict ‘progression’ from infection to disease was identified as another use case, with strong agreement in the survey (89%) and consensus meeting. Identifying individuals at high risk of progressing from infection to disease would allow targeting interventions such as TPT to those most likely to benefit, reduce TB disease incidence and transmission, and improve efficiency of prevention programmes. Interest-holders noted that progression may be influenced by external variables that change with time (e.g. nutrition, aging, co-morbidities, immunosuppressive drugs), and therefore progression tests may need to be repeated.

#### TB disease detection care pathway stage

TB disease is defined as having macroscopic pathology due to *Mtb*, with or without clinical signs or symptoms.^[20]^ Four use cases related to the detection of TB disease were included in the final framework (**Fig 2**).

##### Can we ‘rule out’ TB disease?

One use case focused on ‘ruling-out’ TB disease (93% agreement through survey), i.e. a test with high sensitivity, and therefore a high negative predictive value for excluding TB disease among high-risk populations being screened for TB disease or individuals with presumed TB disease requiring triage. Accurate rule-out would strengthen systematic screening strategies and reduce the burden and cost of microbiological testing for TB disease confirmation.

##### Is there evidence of TB disease?

A second use case related to evidence of or ‘confirmation’ of TB disease (99% agreement through survey). This encompasses rapid, accurate diagnostics for pulmonary and extrapulmonary TB, including non-sputum-based tools and tests suitable for specific populations such as children and people living with HIV. Early and accurate diagnosis can reduce morbidity, mortality, and catastrophic costs, and support transmission reduction. It was noted that a biomarker or test for the detection of asymptomatic TB was included in this use case, but testing for asymptomatic TB disease would likely occur in the context of screening as individuals with asymptomatic disease are less likely to present to health care facilities.

##### Is TB disease infectious?

Assessment of infectiousness fell between 70-80% agreement in the survey (77%) but was ultimately retained as a use case related to TB disease detection (30/34 [88%] of interest-holders polled voted to include) after interest-holder feedback and discussions highlighted the potential patient-level benefits of determining when an individual is no longer infectious, including reduced stigma and earlier return to community life. From a public health perspective, better tools to assess infectiousness could refine isolation practices and transmission control.

##### Is there evidence of drug-resistant TB disease?

Detection of drug-resistant TB disease was agreed as a use case with strong agreement across interest-holder groups (96% agreement through survey). Rapid identification of resistance supports appropriate regimen selection, reduces treatment failure and amplification of resistance, and curtails transmission of drug-resistant TB.

##### Does asymptomatic TB require treatment?

A further, separate use case described whether asymptomatic TB requires treatment (e.g. to prevent progression to symptomatic disease or transmission to contacts). However, interest-holders rejected this use case both through the survey (69% agreement) and in the consensus meeting (25/37 [68%] of interest-holders voted to exclude) due to concerns that any confirmed TB disease warrants treatment and determining duration or type of treatment is separately covered under the treatment stratification use cases.

#### Treatment of TB disease care pathway stage

TB disease treatment was separated from the TB detection stage for the purpose of use case classification and considers the point from which TB disease is diagnosed through to completion of treatment. Three use cases related to treatment optimisation and assessment of treatment response were identified (**Fig 2**).

##### Does this individual need a more intensive TB treatment regimen to achieve a good outcome?

The use case ‘baseline treatment stratification’ seeks to identify individuals who require more intensive or prolonged regimens (i.e. compared to standard or shortest efficacious treatment regimens),^[26]^ or conversely those who could be treated with shorter or less intensive regimens.^[21]^ Such tests could reduce pill burden and side-effects, while maintaining high cure rates, and optimise resource allocation for TB treatment programmes. It received 93% agreement in the survey. It was highlighted that there was good evidence that most individuals could be cured with shorter (e.g. 4-month) regimens, but better tools were needed to predict who these individuals are.

##### Is there inadequate response to TB treatment or risk of poor outcome with current treatment?

The use case for ‘on-treatment monitoring’ addresses early identification of inadequate responses or risk of poor outcomes during therapy (88% agreement through survey). Biomarkers indicating microbiological clearance or risk of poor outcome could enable timely regimen adjustment and reduce relapse and transmission.

##### Is this individual ‘cured’ at the end of their TB treatment, with low risk of relapse?

A further use case concerned ‘confirmation of cure at treatment completion’, identifying individuals at residual risk of relapse (93% agreement through survey). Reliable ‘tests-of-cure’ could provide reassurance to patients and enable targeted follow-up for those at higher risk, improving long-term outcomes. Interest-holders highlighted that all treatment-related use cases could have significant psychosocial implications for patients, for whom TB treatment can often be a challenging and protracted process ^[27]^.

#### Post-TB care pathway stage

Use cases at end of treatment and in post-TB care pertain to tests which provide actionable information after an individual is determined to have achieved a good treatment outcome, which is considered to be bacteriological or clinical improvement, or both, at the end of treatment for TB without evidence of relapse within 6 months.

##### Is there evidence of or risk for post-TB lung disease?

The final use case relates to detection or risk of post-TB lung disease following treatment completion (**Fig 2**). Although biological heterogeneity was acknowledged, interest-holders agreed that tools identifying individuals at risk of post-disease morbidity, ideally before or at treatment completion, could enable monitoring and early intervention to mitigate chronic impairment, which is a significant issue among TB survivors (83% agreement through survey). At the population level, improved recognition and management of post-TB sequelae could reduce long-term health and societal impacts.

##### Is recurrence of disease relapse or re-infection?

A potential use case considering whether recurrence of TB was due to ‘relapse or re-infection’ was carried forward to discussion at the consensus meeting (79% agreement through survey) but was ultimately excluded by interest-holders (27/38 [71%] of interest-holders polled voting to exclude) due to limited clinical consequence, as the current management approach centres on treating recurrent TB and any associated drug resistance regardless of the whether it was newly acquired infection or relapse. Some interest-holders indicated that it may be of greater value for the research community than for public health and clinical utility (e.g., as an end point in vaccine research).

### Prioritisation of use cases

Across the TB spectrum of infection and disease, interest-holders indicated that they prioritised use cases with the greatest potential to reduce mortality, incidence, and transmission in high-burden settings. Detection of drug resistance and prediction of progression from *Mtb* infection to TB disease were ranked as highest importance overall and amongst most interest-holder groups (**Table 2**), followed by progression, evidence of disease and current *Mtb* infection. Use cases related to monitoring responses to TB treatment and treatment stratification were considered moderate priority by most interest-holders. Lower-ranked use cases were those perceived as less actionable, of lower public health impact, or already addressed by existing tools. Use cases that had lower agreement also ranked low in priority, with immune memory ranking the lowest. Differences in prioritisation amongst interest-holder groups (**S11 Table)** were explored in-depth at the consensus meeting.

**Table 2:**
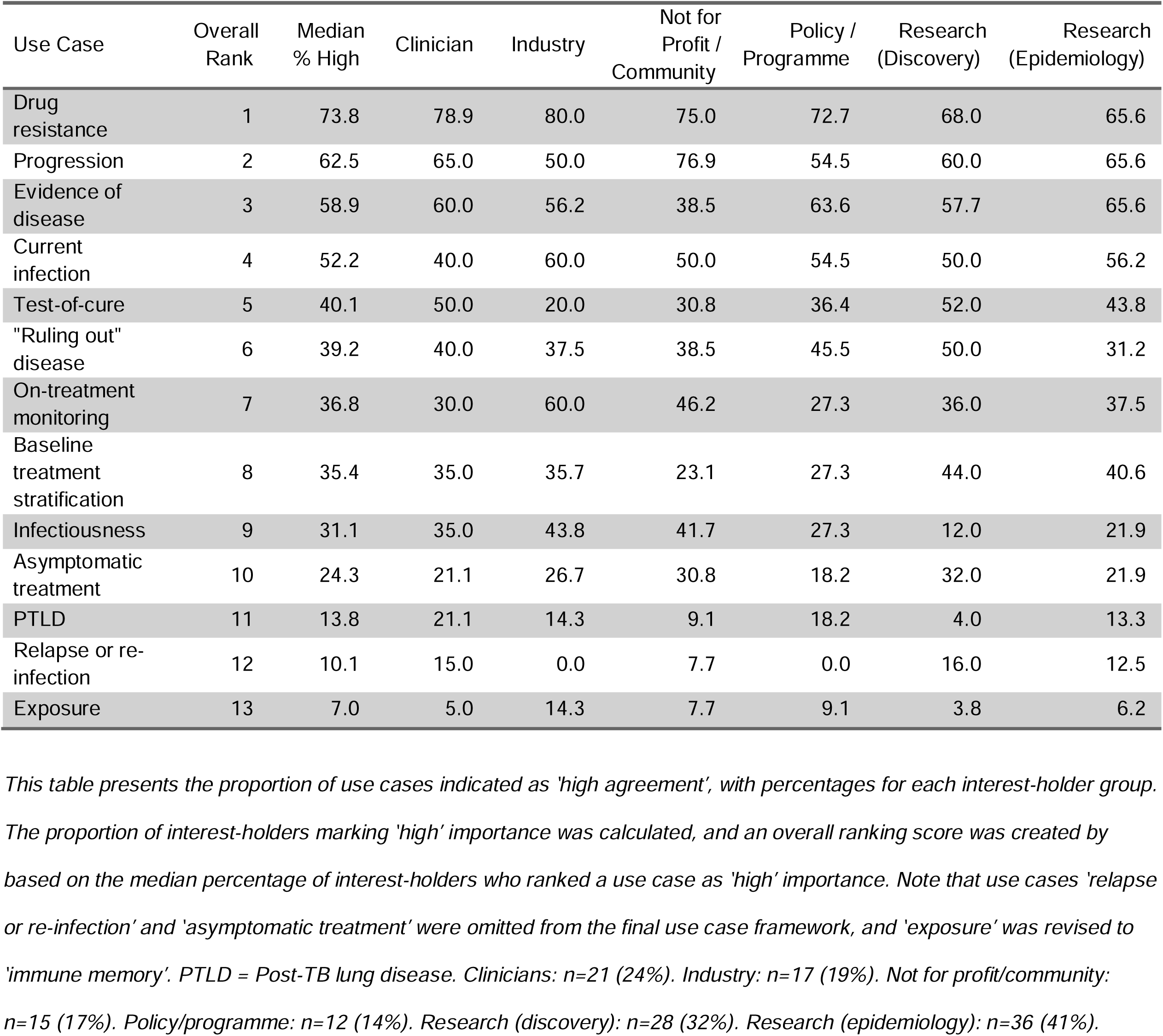
Use cases allocated “high importance” by percentage across interest-holder groups.

| Use Case | Overall Rank | Median % High | Clinician | Industry | Not for Profit / Community | Policy / Programme | Research (Discovery) | Research (Epidemiology) |
| --- | --- | --- | --- | --- | --- | --- | --- | --- |
| Drug resistance | 1 | 73.8 | 78.9 | 80.0 | 75.0 | 72.7 | 68.0 | 65.6 |
| Progression | 2 | 62.5 | 65.0 | 50.0 | 76.9 | 54.5 | 60.0 | 65.6 |
| Evidence of disease | 3 | 58.9 | 60.0 | 56.2 | 38.5 | 63.6 | 57.7 | 65.6 |
| Current infection | 4 | 52.2 | 40.0 | 60.0 | 50.0 | 54.5 | 50.0 | 56.2 |
| Test-of-cure | 5 | 40.1 | 50.0 | 20.0 | 30.8 | 36.4 | 52.0 | 43.8 |
| "Ruling out" disease | 6 | 39.2 | 40.0 | 37.5 | 38.5 | 45.5 | 50.0 | 31.2 |
| On-treatment monitoring | 7 | 36.8 | 30.0 | 60.0 | 46.2 | 27.3 | 36.0 | 37.5 |
| Baseline treatment stratification | 8 | 35.4 | 35.0 | 35.7 | 23.1 | 27.3 | 44.0 | 40.6 |
| Infectiousness | 9 | 31.1 | 35.0 | 43.8 | 41.7 | 27.3 | 12.0 | 21.9 |
| Asymptomatic treatment | 10 | 24.3 | 21.1 | 26.7 | 30.8 | 18.2 | 32.0 | 21.9 |
| PTLD | 11 | 13.8 | 21.1 | 14.3 | 9.1 | 18.2 | 4.0 | 13.3 |
| Relapse or re-infection | 12 | 10.1 | 15.0 | 0.0 | 7.7 | 0.0 | 16.0 | 12.5 |
| Exposure | 13 | 7.0 | 5.0 | 14.3 | 7.7 | 9.1 | 3.8 | 6.2 |

This table presents the proportion of use cases indicated as 'high agreement', with percentages for each interest-holder group. The proportion of interest-holders marking 'high' importance was calculated, and an overall ranking score was created by based on the median percentage of interest-holders who ranked a use case as 'high' importance. Note that use cases 'relapse or re-infection' and 'asymptomatic treatment' were omitted from the final use case framework, and 'exposure' was revised to 'immune memory'. PTLD = Post-TB lung disease. Clinicians: n=21 (24%). Industry: n=17 (19%). Not for profit/community: n=15 (17%). Policy/programme: n=12 (14%). Research (discovery): n=28 (32%). Research (epidemiology): n=36 (41%).

The final TB use case framework (**Fig 2)** integrates pathophysiological distinctions along the TB spectrum with pragmatic decision points in the care of TB programmes, providing a structured foundation to align biomarker research and development with clearly defined clinical and public health objecti

## Discussion

Despite decades of biomarker and test discovery research, there remain significant diagnostic gaps across the spectrum of *Mtb* infection and TB disease that will continue to impede progress towards global TB elimination targets.^[1]^ To our knowledge, this is the first effort to comprehensively identify, consolidate, validate, and prioritise use cases across the entire TB spectrum in a single, consensus-driven framework. By bringing these use cases together, we aim to provide clarity for developers, academics, funders, and policymakers seeking to align innovation with clinical and public health impact.

A central premise of this work is that biomarker development must be grounded in clearly defined, consensus-based, and actionable use cases.^[28]^ Without this, promising biomarkers being developed will not have a clear pathway to implementation or impact. Importantly, we deliberately sought input from a broad range of interest-holders, including industry and test developers, clinicians, programme implementers, policymakers, community representatives, and people with lived experience of TB. This diversity was critical, and the variety of perspectives had significant implications for the final framework. Industry interest-holders highlighted feasibility, scalability, and market considerations; researchers promoted the scientific value and long-term potential, particularly for TB vaccines research; clinicians emphasised immediate patient benefit and reduction in mortality; programme implementers focused on operational fit and equity; and community representatives underscored stigma, acceptability, and quality of life. Incorporating these perspectives will strengthen the validity and relevance of the framework and ensure that biomarker and test development is not driven solely by academic or commercial interests, and acceptability of biomarkers and tests amongst end-users and those affected by TB.^[29]^

The prioritisation exercise revealed a clear pattern: interest-holders assigned highest priority to those use cases that address major gaps in current diagnostic and programmatic practice, and that have the greatest potential to substantially reduce TB mortality, incidence, and transmission. In particular, identification of drug-resistant TB disease, prediction of progression from *Mtb* infection to TB disease, and detection of current, viable *Mtb* infection were consistently prioritised. Similarly, highly sensitive rule-out tests and predictive biomarkers for progression were seen as potentially transformative for scaling preventive therapy and interrupting transmission. At the same time, interest-holders recognised that several of these high-priority use cases are biologically and technically challenging.

Our findings mirror those of the consensus process undertaken by WHO in 2014 to identify high priority TPPs for new tuberculosis diagnostics. However, drug sensitivity testing emerged as a higher priority in our consensus process, which may signal a shift in the field towards use cases beyond screening and detection. It also may reflect a shortcoming of our method, which didn’t explicitly ask survey respondents to ‘rank’ certain use cases above others, but rather identified which use cases were consistently determined to be high priority.

Nonetheless, articulating these priorities explicitly is important, as it signals to researchers and funders where innovation is most urgently needed and where incremental improvements to existing tools may be insufficient. While all identified use cases have potential application in different settings or subpopulations, rapid development and equitable implementation of biomarkers or tests to address the most important use cases are essential to reducing the impact and burden of TB. Importantly, discussions also highlighted where current tools, such as immunosensitisation assays, are not adequate to address key use cases and thus may not merit sustained attention.

Our findings complement, but do not duplicate or replace, the extensive deliberations that have informed TB TPPs from the WHO and other research consortia.^[13,30]^ TPPs have rigorously defined desired characteristics for tests within specific domains, including screening,^[31]^ diagnosis and drug susceptibility testing,^[32]^ or treatment monitoring. ^[21]^ However, this is the first time use cases have been systematically reviewed across all TB stages and then prioritised alongside one another. This cross-spectrum perspective allows comparison of relative importance and highlights trade-offs in resource allocation for biomarker discovery and validation. To build evidence that is interpretable and comparable between studies for each use case, there is a need for clear guidance around evidence generation. Previous guidance has focused on, for example, non-sputum based biomarkers and treatment monitoring tests,^[33,34]^ with forthcoming guidance on TB disease detection to be published by WHO in 2026.^[35]^ Evidence generation guidance for other highly ranked use cases, e.g., ‘progression’, should be prioritised.

Strengths of this work include the systematic scoping review underpinning the framework, the large, comprehensive and geographically diverse interest-holder group, explicit inclusion of community and industry perspectives, and transparent prioritisation methodology. The iterative consensus process allowed contentious use cases to be debated and refined rather than included or excluded uncritically, acknowledging the collective expertise of the TB community and complementing other ongoing interest-holder engagement work.^[4]^ This framework approach could also be used to define use cases for other infections or diseases, especially those with relatively limited research and development funding.

This work also has limitations. We conducted only one formal round of Delphi-survey prior to interest-holder meetings. This was justified by the high degree of consensus achieved in the first round and by a desire to minimise respondent fatigue, which can undermine engagement and diversity of participation.^[16,17]^ Importantly, findings were subsequently validated and refined in two meetings, partially mitigating this limitation. A second limitation is the relatively high proportion of researchers among participants. However, these researchers represented diverse disciplines, including discovery science, epidemiology, clinical research, and implementation, and prioritisation results were stratified by group to account for differing representation. We also recognise the high participation of researchers as an expression of the need for more efforts in advocacy and mobilisation in TB biomarker and test research across broader interest-holders. Representation from high TB burden settings and low- and middle-income countries was strong, enhancing the global relevance of the findings.

A next step is to apply this prioritised use case framework to the current landscape and pipeline of biomarkers and tests currently in development. Doing so will allow identification of mismatches between innovation pipelines and high-priority needs. Beyond scientific discovery, additional barriers to translation, including regulatory pathways, manufacturing capacity, health system readiness, and financing, must also be addressed. Importantly, funders must focus resources towards large, representative and adaptive studies for priority use cases. This use case framework should be considered dynamic and subject to ongoing review as evidence, technologies, and programmatic priorities evolve. Although articulating the use cases is an essential step, ensuring current and future tools get to the right people in an equitable and impactful fashion remains the key challenge for all use cases.^[4]^

In summary, we describe a consolidated and prioritised set of use cases for TB biomarkers and tests derived through a multi-step, interest-holder informed consensus process. By clarifying where biomarkers and tests can most meaningfully influence patient outcomes and public health impact, this framework provides a roadmap for more strategic innovation. The challenge now lies with the TB research, industry, and funding communities to discover, validate, and translate tools that address these high-priority use cases, optimising outcomes for people with *Mtb* infection and TB disease, reducing transmission, and accelerating progress towards ending TB as a public health problem.

## Data Availability

No data was generated by this study.

## Additional information

## Acknowledgments

The authors would like to acknowledge the contribution of all the interest-holders involved in the TB-BIOATLAS project.

## Supporting information

**S1 Table.** Search strategy.

**S2 Table.** Scoping review inclusion criteria.

**S3 Table.** Scoping review exclusion criteria.

**S4 Table.** Data extraction form for scoping review.

**S5 Table.** Interest-holder mapping and rationale for inclusion in the project.

**S6 Appendix.** Survey questions posed to interest-holders in consensus process.

**S1 Figure.** PRISMA 2020 flow diagram for new systematic reviews which included searches of databases and registers only.

**S7 Table.** List of articles included in scoping review.

**S8 Table.** Use cases and sub-use cases for TB biomarkers and tests found in scoping review.

**S9 Table.** Interest-holder groups and demographics of survey respondents and first interest-holder meeting attendees.

**S2 Figure.** Interest-holder survey agreement for all use cases.

**S10 Table.** Summary of survey comments, key messages from first interest-holder meeting, polling outcome, and ultimate decision to retain or omit use cases.

**S11 Table.** The percentage of interest-holders in each group who agreed with the use cases.

