## Supplemental Materials for "Defining use cases for biomarkers and tests across tuberculosis infection, disease and treatment: An international consensus and prioritisation exercise"

### S1 Table: Search strategy.

The following search terms were used for this scoping review search (search date 21/08/2025):

| **Databases** | **Platform** | **Search Strategy (Copy/Paste)** | **Limits Applied** |
| --- | --- | --- | --- |
| PubMed / MEDLINE | NCBI (https://pubmed.ncbi.nlm.nih.gov) | ("tuberculosis"[MeSH Terms] OR "tuberculosis"[Title/Abstract]) AND ("target product profile"[Title/Abstract] OR TPP[Title/Abstract] OR "preferred product characteristics"[Title/Abstract] OR PPC[Title/Abstract]) | English language; Humans |
| Global Index Medicus | WHO Global Index MedicusIt re (https://www.globalindexmedicus.net) | (tuberculosis) AND (TW:"target product profile" OR TW:TPP  OR TW:"preferred product characteristics" OR TW:PPC) | English language |
| WHO IRIS | WHO.int (https://iris.who.int) | tuberculosis AND ("target product profile" OR TPP OR "preferred product characteristics" OR PPC) | English language |
| FIND website | Google.com (site:finddx.org) | "tuberculosis" AND ("target product profile" OR TPP OR "preferred product characteristics" OR PPC) | English language |
| WHO website | Google.com (site:who.int) | "tuberculosis" AND ("target product profile" OR TPP OR "preferred product characteristics" OR PPC) | English language |

### S2 Table: Scoping review inclusion criteria.

| Criterion | Description |
| --- | --- |
| **Population** | Studies involving individuals with presumptive TB, Mtb infection or TB disease, or being screening for TB, including all age groups and risk populations (e.g. PLHIV, household contacts, congregate settings, migrants). |
| **Focus** | Documents describing target product profiles (TPPs) or guidelines relating to the management of any state or stage of TB (including extrapulmonary TB), covering steps such as symptom recognition, care-seeking, screening, testing, diagnosis, referral, treatment initiation/monitoring, and post-TB care. |
| **Content** | Must report on Target product profiles (TPPs) and at least one of the following:  – Regional, international, or global guidelines  – WHO-endorsed tools, diagnostics, or treatment approaches in clinical use  – Systematic reviews related to TB TPPs or guidelines. |
| **Study type** | Quantitative, qualitative, mixed-methods, implementation, modelling, or operational research studies, program evaluations are eligible. |
| **Geography** | Regional, international, or global sources. National or subnational level documents excluded. Emphasis on high-TB burden settings. |
| **Language** | Articles in English (or those with non-English abstracts if data extraction is feasible through AI-translation tools). |
| **Publication date** | Any year; for updated guidelines, only the most recent version included unless older versions contained unique details not reproduced in newer ones. |

### S3 Table: Scoping review exclusion criteria.

| **Criterion** | **Description** |
| --- | --- |
| **Focus not on TPPs** | Studies that do not describe or explicitly refer to target product profiles (TPPs) |
| **No relevance to TB** | Studies focused on other diseases without integrated Mtb infection or disease care components. |
| **No insight into use cases, decision points or care gaps** | Publications that do not provide practical or applicable information for TB care pathways or product development. |
| **Commentaries, editorials, or protocols only** | Opinion pieces, narrative commentaries, editorials, or study protocols not presenting empirical data or finalized frameworks. |
| **National/subnational scope only** | Documents pertaining solely to country- or subnational-level guidelines (excluded due to heterogeneity among NTPs). |
| **Older versions of updated guidelines** | When superseded by more recent WHO or international guidelines, unless unique details are only available in earlier versions. |
| **Unavailable full text** | If the abstract does not contain sufficient information and full text cannot be retrieved. |

### S4 Table: Data extraction form for scoping review.

| 1. **Key** |
| --- |
| 2. **Reviewer name** |
| 3. **Use Case ID** |
| 4. **Source Document** |
| 5. **Author(s)** |
| 6. **Year published** |
| 7. **Use case scenario description** |
| 8. **Target population** |
| 9. **Mapped TB Disease State** |
| 10. **Care pathway step(s)** |
| 11. **Potential patient-level outcomes and** |
| 12. **Potential population-level outcomes and benefits** |
| 13. **Examples of other tests or biomarkers for this use case.** |
| 14. **Setting - Selected Choice** |
| 15. **Operational requirements** |
| 16. **Intended end-user - Selected Choice** |
| 17. **Outcome of test** |
| 18. **Timing** |
| 19. **Price** |
| 20. **Any other notes** |

### S5 Table: Interest-holder mapping and rationale for inclusion in the project.

| **Interest-holder group** | **Rationale for inclusion** | **Example groups, organisations or consortia** |
| --- | --- | --- |
| **Researchers (Discovery)** | Contribute perspective on discovery science, early validation and generating evidence towards biomarkers and tests for different use cases | Academic research groups focused on discovery of host-responses to MTB and pathogen-derived biomarkers, including with a mechanistic focus |
| **Researchers (Epidemiology / Clinical)** | Provide insights into clinical validation and clinical impact studies of tests or biomarkers, broader perspectives of epidemiology and determinants of TB in target populations, and the potential impact of findings/tests for identified use cases | Consortia and groups focused on high-quality clinical validation and large-scale trials in representative subpopulations. These include diagnostic evaluation studies (e.g. R2D2 TB Network, RePORT) and clinical trial consortia evaluating therapeutics (e.g. UNITE4TB, GoFAST, RADIO-TB) |
| **Industry / diagnostic Developers** | Inform technical feasibility and innovation potential for tests; understand scalability, commercialisation of biomarkers or tests, markets and regulatory pathways | Commercial entities, small and medium enterprises and technology partners |
| **Clinicians / Field Practitioners** | Interpret clinical utility and feasibility of use cases in real-world settings, offer perspectives on decisions needed to be made in clinical care. | Frontline medical practitioners who frequently treat people with presumed or confirmed Mtb infection and TB disease. |
| **Policymakers / Programme Implementers** | Align use cases with local, regional, national and/or global TB strategies; evaluate operational feasibility and integration into existing health systems, understand the priorities and challenges of TB programmes and broader health systems | International and national health bodies such as:   - WHO (specifically prequalification and target product profile units) - Foundation for Innovative New Diagnostics (FIND) - National TB Programs (NTPs) |
| **Not-for-profit organisations / Community members / People with Lived Experience of TB** | Share lived experience insights; validate relevance and feasibility of use cases; reflect priorities of not-for-profits and community advocacy groups | Global advocacy organisations, such as:   - Stop TB Partnership - The International Union Against Tuberculosis and Lung Disease (The Union) - TB Alliance - Friends for International Tuberculosis Relief - Treatment Action Group (TAG) - TBpeople - Gates Medical Research Institute |
| **Funders** | Provide insights into donor and funder priorities | Funding/intermediary organisations, such as:   - Gates Foundation - National Institute of Health (NIH) - TB-REACH - Wellcome Trust - Unitaid - Global Fund |

| S6 Appendix: Survey questions posed to interest-holders in consensus process. | |
| --- | --- |
| 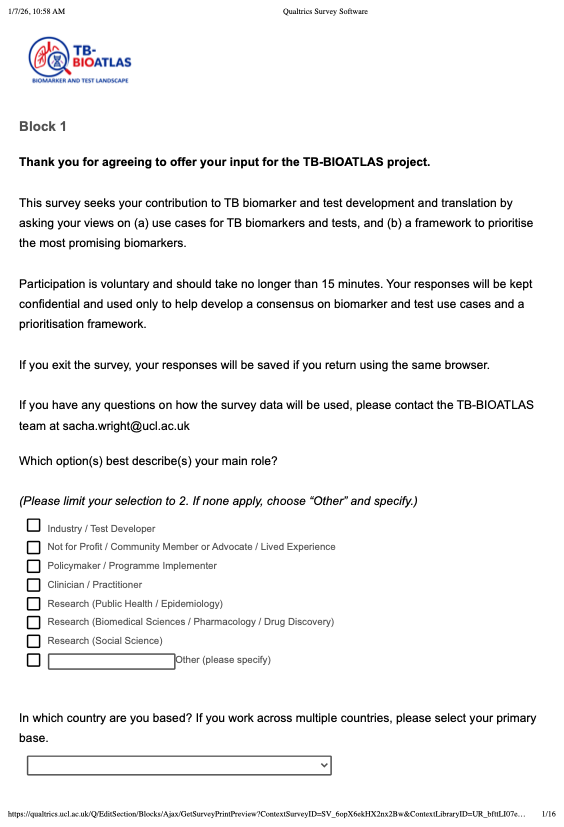 | 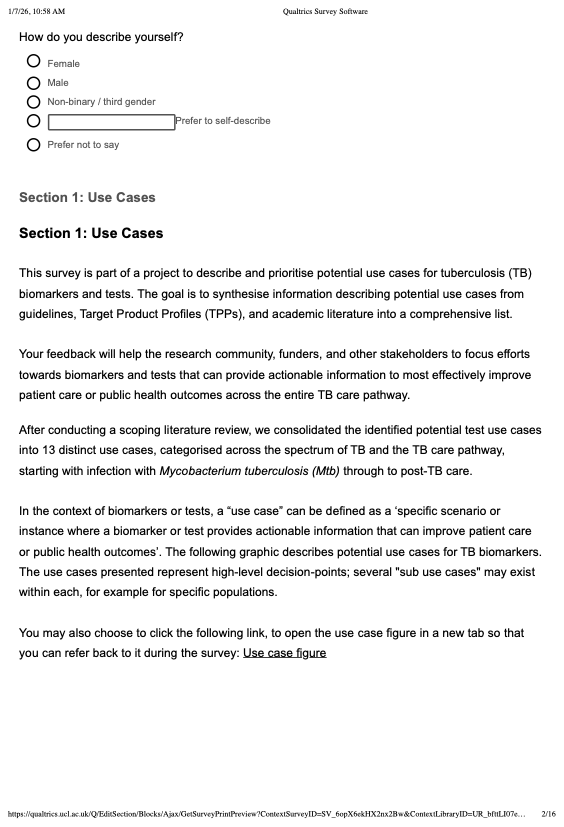 |
| 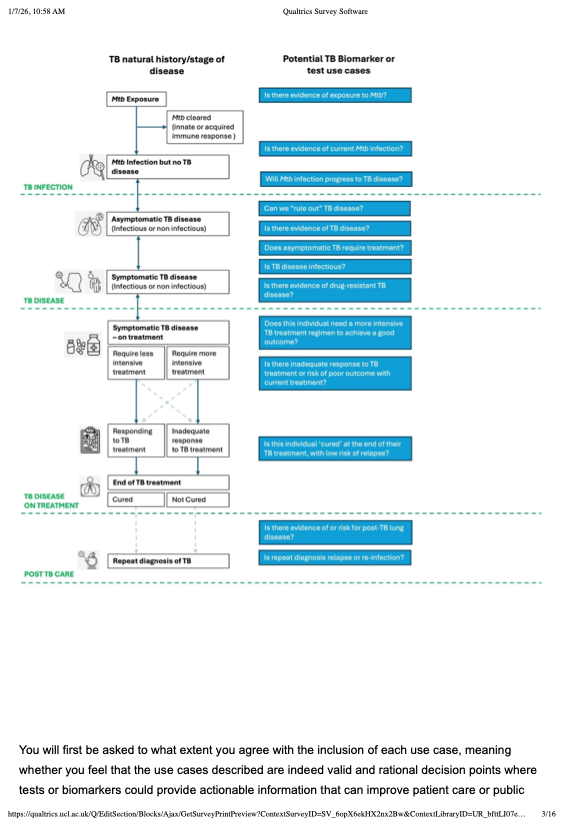 | 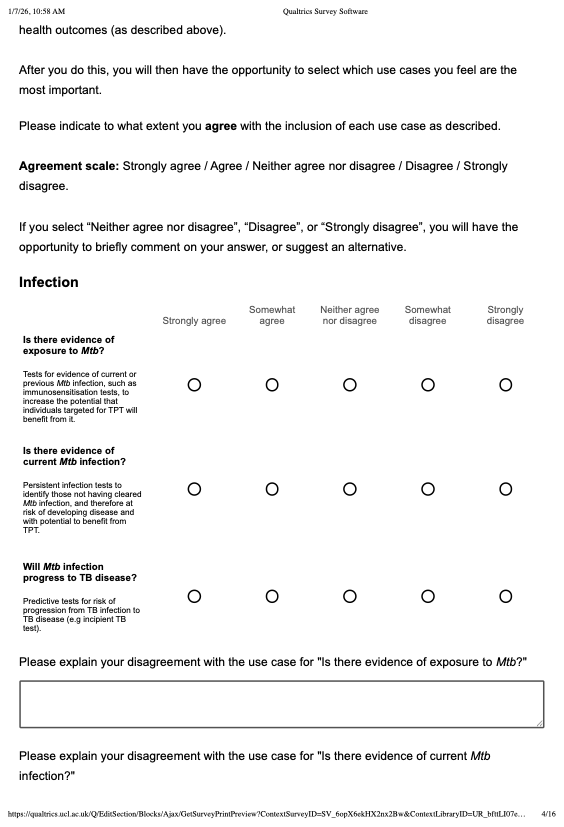 |
| 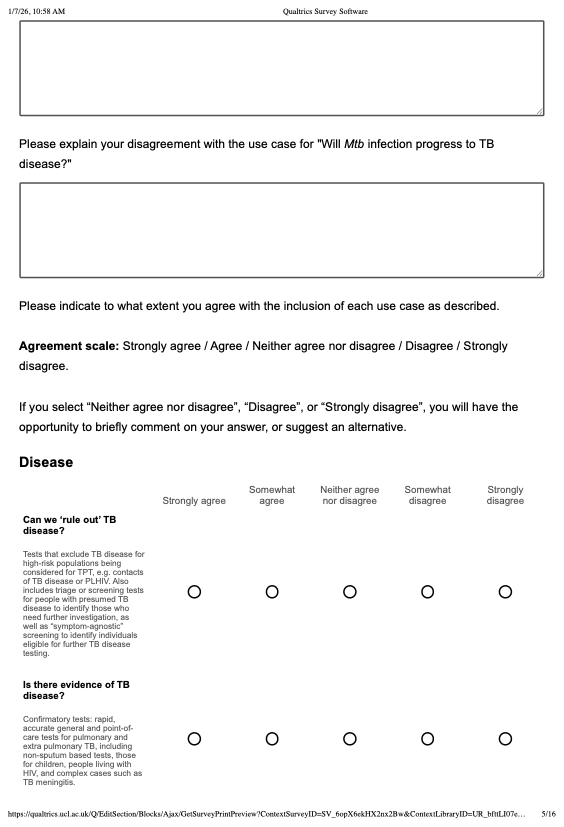 | 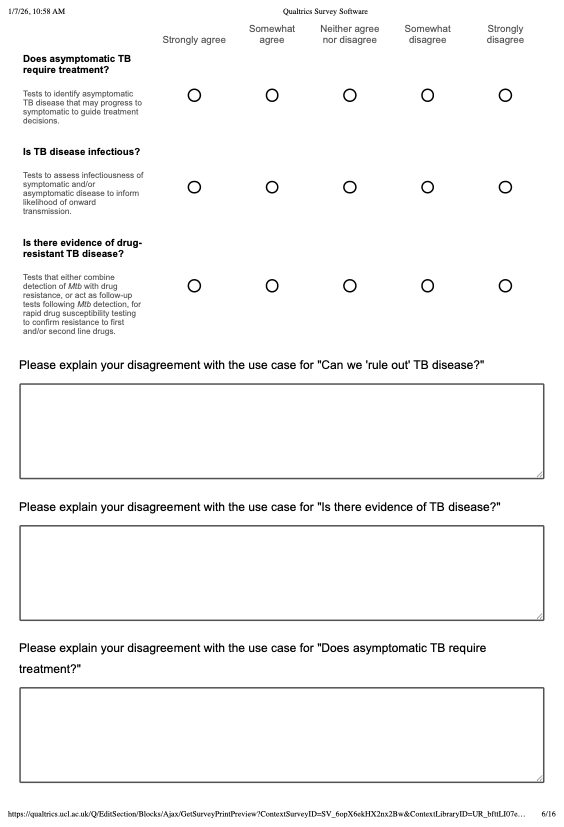 |
| 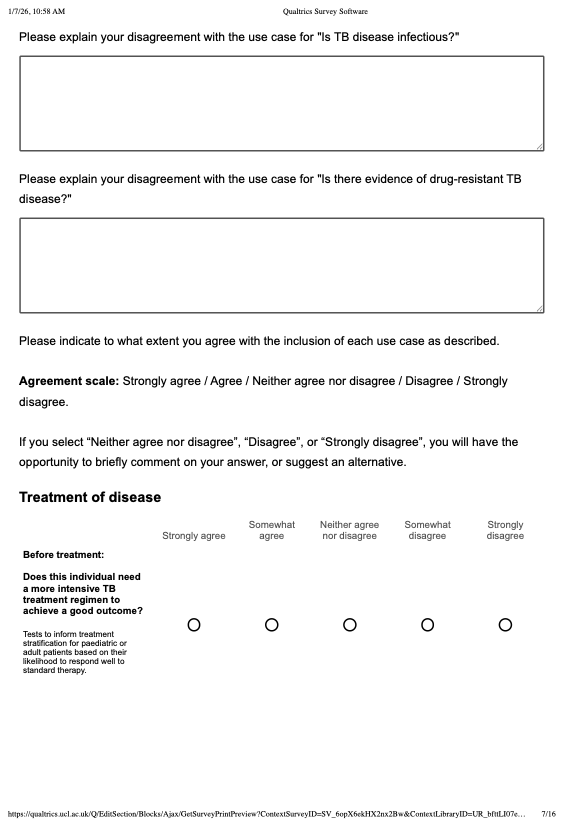 | 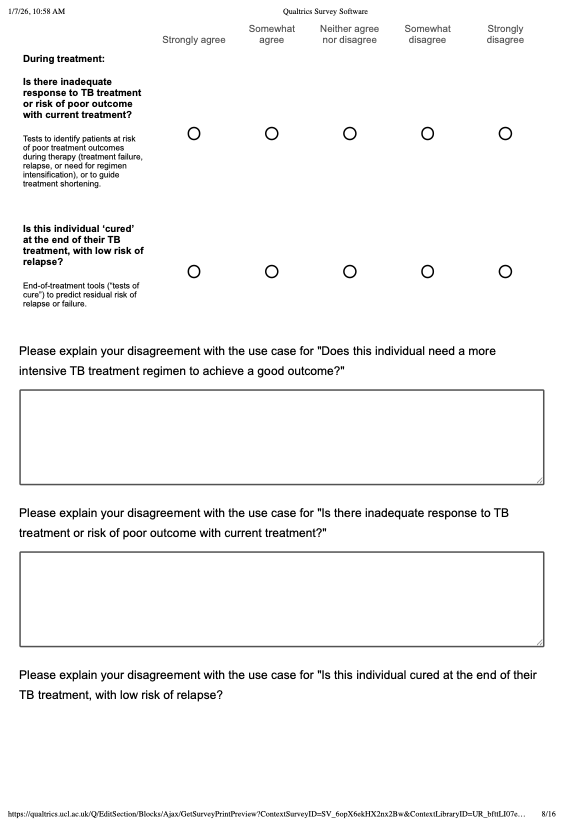 |
| 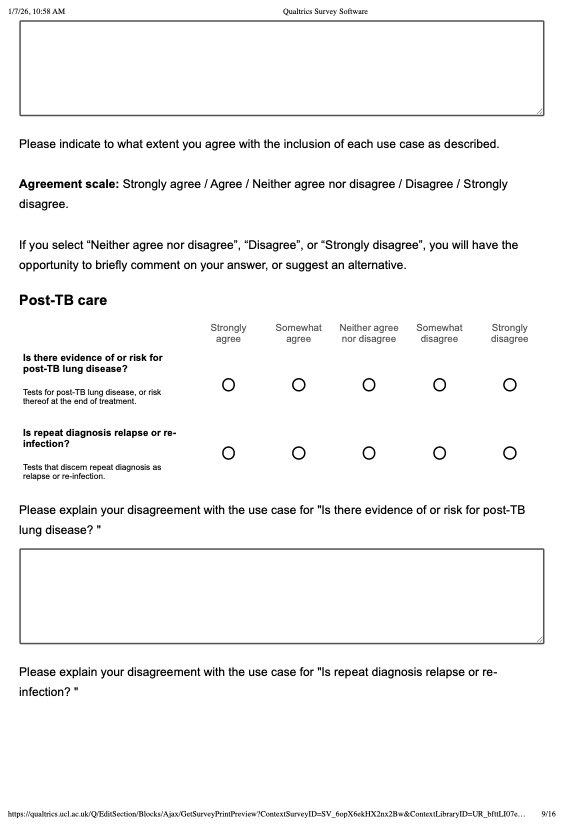 | 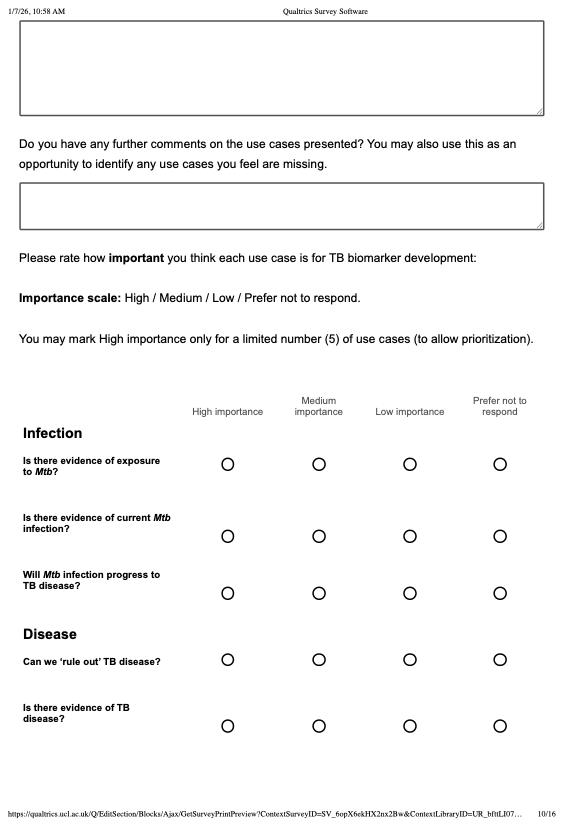 |
| 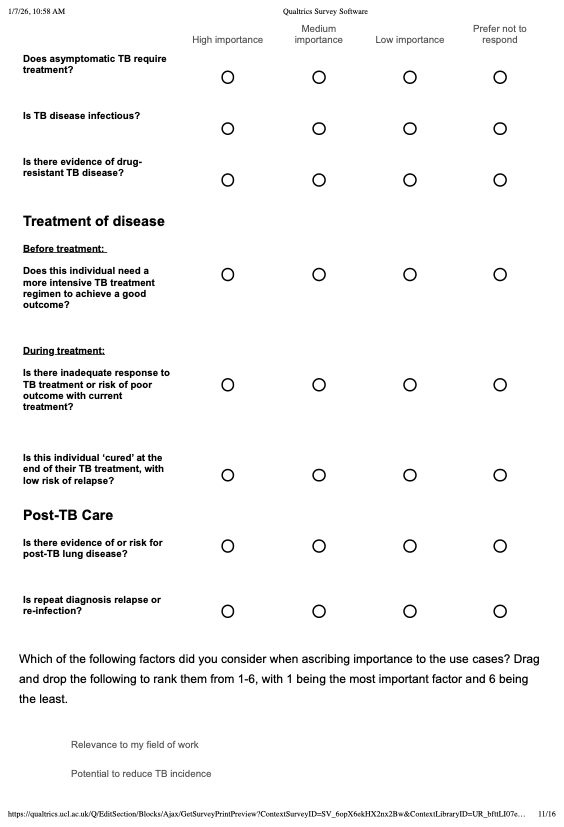 | 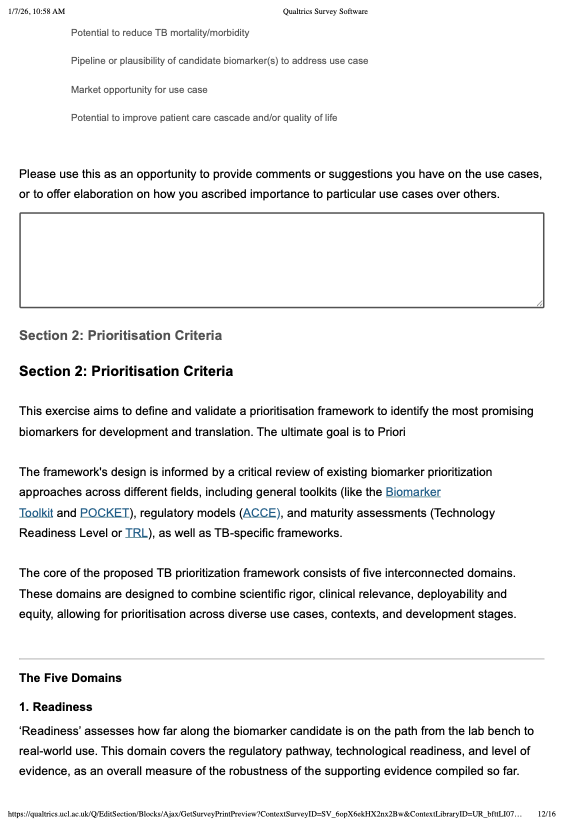 |
| 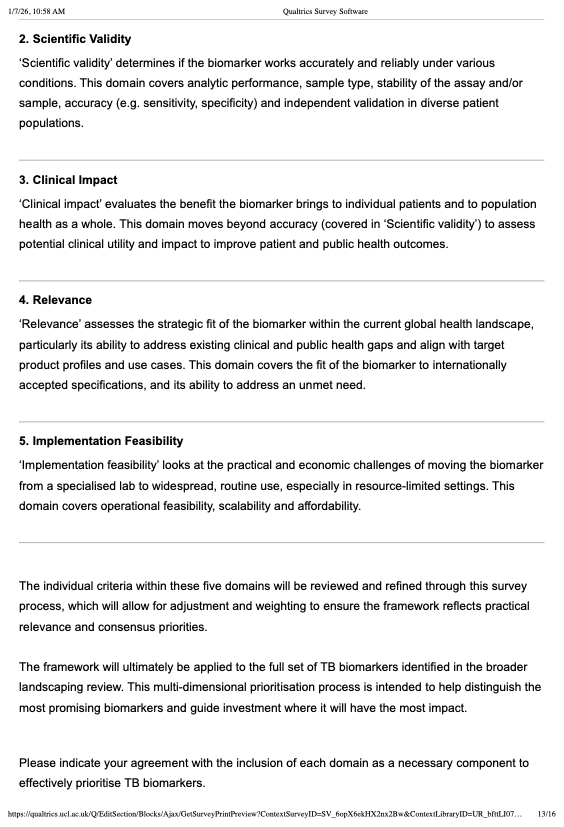 | 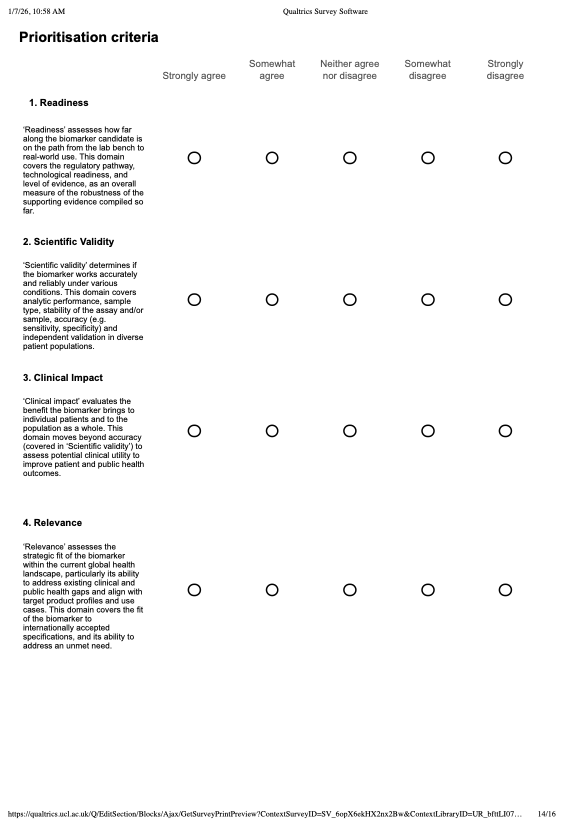 |
| 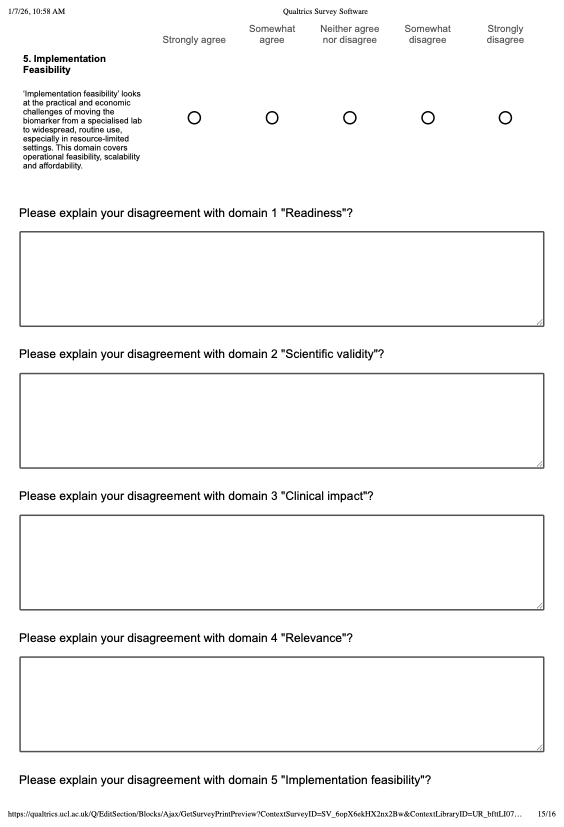 | 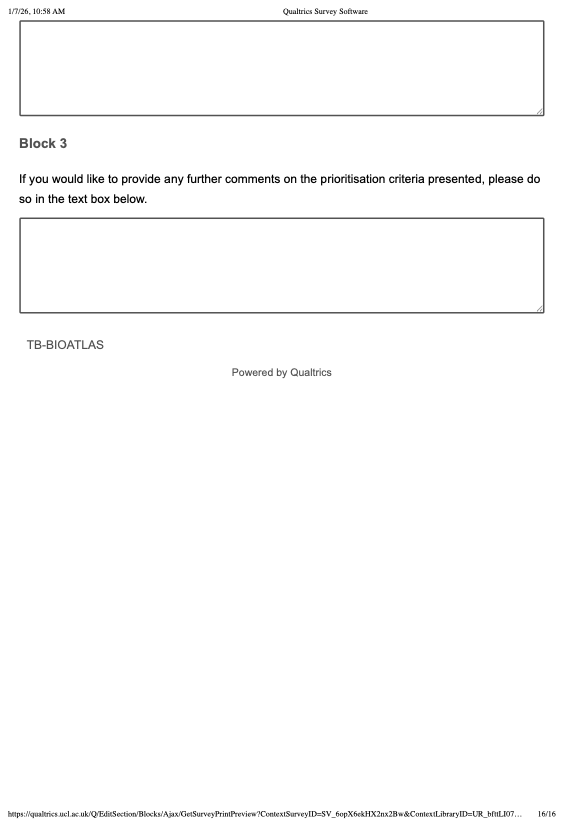 |

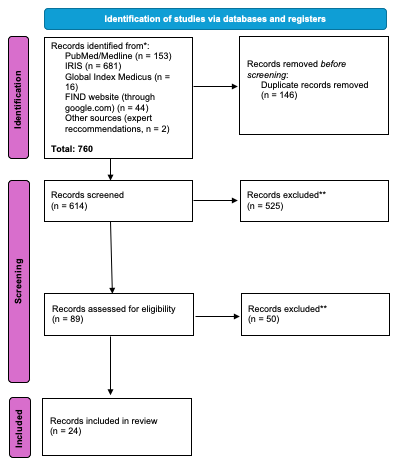
S1 Figure: PRISMA 2020 flow diagram for new systematic reviews which included searches of databases and registers only.

Source: Page MJ, et al. BMJ 2021;372:n71. doi: 10.1136/bmj.n71. This work is licensed under CC BY 4.0.

### S7 Table: List of articles included in scoping review.

| Key | Year | Title | Author | Type of document |
| --- | --- | --- | --- | --- |
| 1 | 2025 | Target product profiles for tuberculosis screening tests | World Health Organization | Target product profile |
| 2 | 2025 | WHO consolidated guidelines on tuberculosis: module 4: treatment and care | World Health Organization | Guideline |
| 3 | 2025 | Rapid tuberculosis diagnosis from respiratory or blood samples by a low cost, portable lab-in-tube assay | Youngquist et al. | Research paper (Science Translational Medicine) |
| 4 | 2025 | Novel and optimized diagnostics for pediatric TB in endemic countries: NOD-pedFEND study protocol (BMC Pediatrics) | Song et al. | Published Research Protocol (BMC Pediatrics) |
| 5 | 2024 | Target product profiles for tuberculosis diagnosis and detection of drug resistance | World Health Organization | Target product profile |
| 6 | 2024 | WHO operational handbook on tuberculosis: module 1: prevention: tuberculosis preventive treatment | World Health Organization | Guideline |
| 7 | 2024 | WHO consolidated guidelines on tuberculosis: module 1: prevention: tuberculosis preventive treatment (2^nd^ edition) | World Health Organization | Guideline |
| 8 | 2024 | WHO operational handbook on tuberculosis: module 3: diagnosis: rapid diagnostics for tuberculosis detection | World Health Organization | Guideline |
| 9 | 2024 | WHO consolidated guidelines on tuberculosis: module 3: diagnosis | World Health Organization | Guideline |
| 10 | 2023 | Evidence and research gaps identified during development of policy guidelines for tuberculosis | World Health Organization | Policy evidence mapping report |
| 11 | 2023 | Target product profiles: tests for tuberculosis treatment monitoring and optimization | World Health Organization | Target product profile |
| 12 | 2022 | WHO consolidated guidelines on tuberculosis: module 5: management of tuberculosis in children and adolescents | World Health Organization | Guideline |
| 13 | 2021 | Target product profile for next-generation drug-susceptibility testing at peripheral centres | World Health Organization | Target product profile |
| 14 | 2021 | WHO consolidated guidelines on tuberculosis: module 2: screening: systematic screening for tuberculosis disease | World Health Organization | Guidelines |
| 15 | 2020 | Target product profiles for tuberculosis preventive treatment | World Health Organization | Target product profile |
| 16 | 2018 | Biomarkers for diagnosis of childhood tuberculosis: A systematic review. | Togun et al. (PLoS One) | Systematic review |
| 17 | 2017 | Consensus Meeting Report: Development of a Target Product Profile (TPP) and a framework for evaluation for a test for predicting progression from tuberculosis infection to active disease | World Health Organization | Target product profile (meeting report) |
| 18 | 2015 | Guidelines for surveillance of drug resistance in tuberculosis 5 th Edition | World Health Organization | Guideline |
| 19 | 2015 | Target product profile of a molecular drug-susceptibility test for use in microscopy centers | Denkinger et al. | Research paper (The Journal of Infectious Diseases) |
| 20 | 2015 | TB Diagnostics Market in Select High-Burden Countries: Current Market and Future Opportunities for Novel Diagnostics | FIND | Financial report |
| 21 | 2014 | High-priority target product profiles for new tuberculosis diagnostics: report of a consensus meeting | World Health Organization | Report (meeting report) |
| 22 | 2013 | Priorities for tuberculosis research: a report of the disease reference group on TB, leprosy and buruli ulcer | WHO, Special Programme for Research and Training in Tropical Diseases (WHO + UNICEF + UNDP + World Bank) | Report |
| 23 | 2025 | Assessment of comorbidities, risk factors, and post tuberculosis lung disease in National Tuberculosis Guidelines: A scoping review | M.S.Bah et al. | Scoping review |
| 24 | 2024 | Classification of early tuberculosis states to guide research for improved care and prevention: an international Delphi consensus exercise | A.K.Coussens et al. | Research paper (Delphi exercise) |

### S8 Table: Use cases and sub-use cases for TB biomarkers and tests found in scoping review.

| TB disease state | Source(s) keys | Main use case   - *Example sub-use cases* | Description of main use case | Shortened title |
| --- | --- | --- | --- | --- |
| *Mtb* infection | 6, 7, 10, 12, 15, 17, 22, 24 | Is there evidence of exposure to *Mtb*? | Tests for evidence of current or previous Mtb infection (e.g. immunosensitisation tests). | Exposure |
|  |  | Is there evidence of current *Mtb* infection?   - *Has Mtb infection re-occurred?* | Persistent infection test to identify those who have not cleared *Mtb* infection, and therefore have detectable levels of *Mtb* and an associated host response, but are without disease. | Evidence of infection |
|  |  | Will *Mtb* infection progress to TB disease? | Predictive tests for risk of progression from current *Mtb* infection (TBI) to active TB disease (also sometimes called an incipient TB test). | Progression |
| TB disease | 1, 3, 4, 5, 6, 7, 8, 10, 12, 14, 15, 16, 19, 20, 22, 24 | Can we ‘rule-out’ TB disease?   - *Can individuals be given TPT for Mtb infection?* - *Does this individual need further (microbiological) testing for TB disease?* | Test that excludes TB disease for high-risk populations being screened for TPT, e.g. contacts of TB disease or people living with HIV (PLWH).  Also includes triage or screening test for people with presumed TB disease to identify those who need further investigation, as well as symptom-agnostic screening to identify individuals eligible for further TB disease testing. | “Ruling-out” disease |
|  |  | Is there evidence of TB disease?   - *Does this individual with signs or symptoms of presumed TB have TB disease?* - *Does this individual have evidence of TB meningitis?* - *Does this asymptomatic individual have TB disease?* | Confirmatory diagnostic tests, preferably rapid, accurate, general and point-of-care tests for pulmonary and extra pulmonary TB, including non-sputum-based tests, those for children, people living with HIV, and complex cases such as TB meningitis. | Evidence of disease |
|  |  | Does asymptomatic TB require treatment? | Tests to identify whether early, asymptomatic TB disease is likely progress to symptomatic disease. | Asymptomatic treatment |
|  |  | Is TB disease infectious? | Test to assess infectiousness of bacteriologically confirmed disease. May also provide information about likelihood of transmission. | Infectiousness |
|  |  | Is there evidence of drug-resistant TB disease?   - *Does this individual require rapid drug susceptibility testing for first- or second-line TB drugs?* - *Does this individual need treatment for drug-resistant TB disease?* - *What TPT regimen is suitable for close contacts of the index case?* | Tests that either combine detection of Mtb with drug resistance, or follow-up tests for rapid drug susceptibility testing to confirm resistance to first and/or second line drugs. | Drug resistance |
| Treatment of TB disease | 2, 8, 9, 10, 11, 12, 13, 18, 21, 22, 24 | Does this individual need a more intensive TB treatment regimen to achieve a good outcome? | Tests to inform baseline treatment stratification for all patients, symptomatic or asymptomatic (paediatric or adult patients, as well as PLWH and complex cases), based on their likelihood to respond well to standard therapy. | Baseline treatment stratification |
|  |  | Is there an inadequate response to TB treatment or risk of poor outcome with current treatment?   - *Has drug resistance developed during the course of treatment?* | Includes biomarkers and tests that can contribute to treatment optimization, which refers to initiating or switching to an effective TB treatment regimen that results in a high likelihood of a good outcome.  This may include tests to monitor treatment response (molecular, microbiological, biomarker-based), tests to identify individuals at risk of poor treatment outcomes during therapy (treatment failure, relapse, or need for regimen intensification), or biomarkers or diagnostics for microbiological clearance and treatment shortening. | On-treatment monitoring |
|  |  | Is this individual ‘cured’ at the end of their TB treatment, with low risk of relapse? | End-of-treatment tools (“test-of-cure”) to identify residual risk of relapse or failure. | Test-of-cure |
| Post-TB care | 10, 18, 23, 24 | Is there evidence of or risk for post-TB lung disease?   - *Is the individual at high risk of developing post-TB lung disease?* | Tests for post-TB lung disease, or risk thereof at end of treatment. | PTLD |
|  |  | Is repeat diagnosis relapse or re-infection? | Tests that discern between repeat diagnosis as recurrence or re-infection. | Relapse or re-infection |

Source(s) keys refer to associated references extracted through the scoping review. Keys are shown in Supplementary Table 7. Where the word “test(s)” is used, it can be assumed this refers to biomarkers or tests. PTLD is Post TB Lung Disease

### S9 Table: Interest-holder groups and demographics of survey respondents and first interest-holder meeting attendees.

|  | **Survey respondents** | **Interest-holder meeting attendees** |
| --- | --- | --- |
| **Interest-holder Characteristics** | **N = 88***^1^* | **N = 48***^1^* |
| **Role** |  |  |
| Clinician | 21 (24%) | 8 (17%) |
| Research (Epidemiology) | 36 (41%) | 9 (19%) |
| Research (Discovery) | 28 (32%) | 14 (29%) |
| Industry | 17 (19%) | 7 (15%) |
| Not for Profit / Community | 15 (17%) | 5 (10%) |
| Policy / Programme | 12 (14%) | 5 (10%) |
| **Gender** |  |  |
| Female | 38 (44%) | 26 (54%) |
| Male | 47 (54%) | 22 (46%) |
| Prefer not to say | 2 (2%) | - |
| Not Specified | 1 | - |
| **WHO Region** |  |  |
| African Region | 24 (27%) | 11 (23%) |
| Eastern Mediterranean Region | 4 (5%) | 3 (6%) |
| European Region | 27 (31%) | 21 (44%) |
| Region of the Americas | 14 (16%) | 8 (17%) |
| South-East Asia Region | 7 (8%) | 0 (0%) |
| Western Pacific Region | 12 (14%) | 5 (10%) |
| **Country Income Level** |  | *Not specified* |
| High Income | 44 (50%) | - |
| LMIC | 44 (50%) | - |
| **Country TB Incidence per 100k** |  | *Not specified* |
| Low to Medium (<40) | 44 (50%) | - |
| High to Very High (≥40) | 44 (50%) | - |
| *^1^*n (%)  WHO = World Health Organization LMIC = Low- and Middle-Income Countries  TB = tuberculosis | |  |

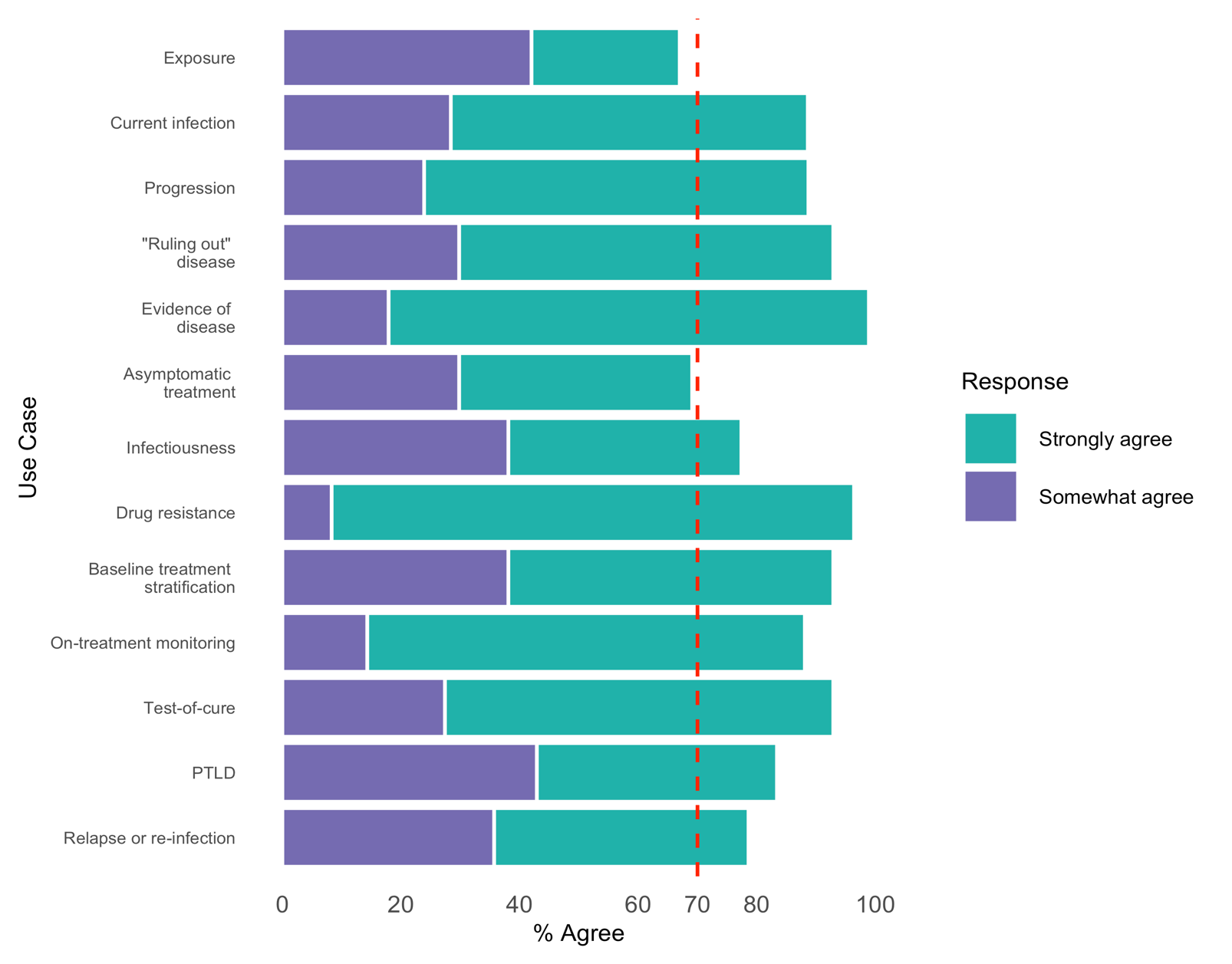

### S2 Figure: Interest-holder survey agreement for all use cases

For each use case, the chart shows the combined percentage of respondents who either ‘Somewhat agree’ (purple) or ‘Strongly agree’ (green). The vertical red line at 70% represents the threshold for consensus. PTLD Post TB Lung Disease

### S10 Table: Summary of survey comments, key messages from first interest-holder meeting, polling outcome, and ultimate decision to retain or omit use cases.

| Use case   (% agreed or strongly agreed) | Summary of survey comments | Interest-holder meeting key messages | Polling outcome | Ultimate decision |
| --- | --- | --- | --- | --- |
| Exposure (67%) | - Respondents stated that they were confused with the difference between the existence of a use case with its priority or importance for reducing TB mortality and morbidity. - In high-burden settings, biomarkers for exposure have low predictive value because nearly everyone is constantly exposed. - Implementation in LMICs may be prohibitively expensive compared to the lower cost of direct treatment or preventive therapy. - There is a significant lack of clinical actionability, as knowing someone was exposed without having an active infection offers little bedside benefit. - The utility of such tests may be limited to surveillance rather than high-priority clinical or public health interventions. | - The term "infection" is currently used too broadly, failing to distinguish between individuals who have successfully cleared the bacteria and those with an active, ongoing infection. - Current diagnostics (like IGRA) measure immune response rather than the presence of viable bacteria; there is a critical need for a test that can confirm "viable infection" directly. - The lack of a reliable infection test is a major bottleneck for vaccine development; without one, clinical trials struggle to measure prevention of infection accurately, leading to wasted resources and misleading data. - In high-stigma environments, testing for exposure carries a high "marginal cost" to the individual, as a positive result can cause significant mental distress or be wrongly equated with having the disease. - Identifying individuals who are exposed but naturally resist infection is a high-priority research goal, as understanding their biological protection mechanisms could unlock new pathways for vaccine design. | Participants voted in the Mentimeter poll 23 to 15 to exclude the use case from the final framework.  Moderators reflected that rejection may stem from poor phrasing and protracted wording debates rather than a dismissal of the use case itself. Interest-holders struggled to distinguish between the existence of a use case and its clinical importance.  TB-BIOATLAS team held a final vote to either remove the use case or retain it with revised language emphasizing its low priority. | Retain, but revise language to “Is there evidence of immune memory to Mtb?” |
| Current *Mtb* infection (89%) | - Infection tests currently used (like IGRAs) detect an immune response rather than the physical presence of bacteria, making them better suited for use cases other than this one (see first use case). - Detecting viable *M. tuberculosis* in the absence of disease likely requires a combination of distinct biomarkers. - A suggested approach involves a two-step testing process: first confirming the presence of viable bacteria, then pairing it with a test to confirm or rule out disease. | N/A |  | Keep in the final framework |
| Progression (89%) | - Progression is driven by external variables (e.g., nutrition, stress, aging, or comorbidities like HIV/diabetes) that may change after a test is administered. - Biomarkers reflect a snapshot of a current state but often lack the predictive power to determine an individual’s future clinical trajectory. - Interest-holders questioned the feasibility of a predictive test due to the low sensitivity and specificity of current assays. | N/A |  | Keep in the final framework |
| Rule-out (93%) | - One comment was raised that this use case seems to cover several use cases. | N/A |  | Keep in the final framework |
| Evidence of disease (99%) | - None. | N/A |  | Keep in the final framework |
| Asymptomatic treatment (69%) | - The lack of a standardized diagnostic definition for asymptomatic TB creates significant ambiguity, particularly regarding bacteriologically unconfirmed cases. - There is strong consensus that any bacteriologically confirmed TB requires treatment to prevent negative outcomes, regardless of whether the individual shows symptoms. - Because confirmed TB is already an automatic trigger for treatment, interest-holders argue that a biomarker to decide whether to treat is unnecessary. - The focus should shift from "if" to treat to "how" to treat, using biomarkers for treatment stratification rather than diagnosis. - Biomarkers could be better utilized for baseline risk assessments to determine the appropriate regimen length and intensity based on bacillary burden and relapse risk. | - Asymptomatic TB is almost exclusively identified through proactive screening rather than patient presentation at clinics. - The asymptomatic label is highly variable and subjective, depending on screening methods, question phrasing, and patient perception. - Technical identification depends heavily on the sensitivity of specific diagnostic tools and sample types, complicating population definitions. - Clinicians view withholding treatment from any bacteriologically confirmed case as ethically problematic, regardless of symptoms. - Clinical priority should shift from deciding whether to treat to determining the most appropriate regimen length and the potential for self-cure. - New biomarkers are needed to identify bacteriologically unconfirmed asymptomatic cases and to serve as correlates of protection for vaccine development. - The asymptomatic use case could be integrated into the broader framework of baseline treatment stratification rather than remaining a standalone category. | Mentimeter polling indicated a clear rejection of the use case, with 25 of 37 respondents voting to omit the use case from the final framework. | Omit from the final framework |
| Infectiousness (77%) | - Confirming a TB diagnosis demands immediate treatment and linkage to care, making the assessment of infectiousness a secondary clinical priority. - A standalone test for infectiousness may have limited real-world utility for public health programs and relies on problematic clinical definitions of symptoms. - Developing a single, reliable test is considered unfeasible because infectivity depends on external variables like ventilation, proximity, and the recipient's susceptibility. - The most valuable application for an infectiousness marker is determining when a individual on treatment can safely end isolation and return to the community. - Interest-holders suggest that transmission potential is dynamic and changes over time, requiring careful description rather than a static measurement. | - Community interest-holders challenged the less important rating from researchers, arguing that previous feedback overlooked the lived experiences of individuals. - A definitive test for non-infectiousness is highly valued across all settings to safely determine the end of isolation. - Such a test would prevent prolonged, unnecessary isolation for DR-TB patients, reducing associated mental and social harm. - Advocates compared the potential impact to the HIV “U=U” principle, noting it could dismantle the social stigma that drives fear and isolation. - Clear information regarding infectiousness is seen as “incredibly empowering,” likely increasing testing demand and improving community health-seeking behavior. - Reliable markers would allow patients to return to their families sooner, removing the fear of transmitting the disease to loved ones. | 30 out of 34 respondents agreed in the Mentimeter poll that this should be included as a use case. | Keep in the final framework |
| Drug resistance (96%) | - Some comments implied skepticism that drug resistance could be detected through a single test. | N/A |  | Keep in the final framework |
| Baseline treatment stratification (93%) | - Interest-holders suggested rephrasing the use case to focus on identifying individuals eligible for shorter, less intensive treatment regimens that achieve non-inferior outcomes. - Some respondents emphasized that differentiated care should be person-centered rather than solely biomarker-driven. - Feedback highlighted the importance of patient autonomy, suggesting individuals should be empowered to participate in decision-making regarding their treatment options. | N/A |  | Keep in the final framework |
| On-treatment monitoring (88%) | - Comments suggested skepticism that a single biomarker could be sufficient; would likely require a combination of multiple markers or factors. - Current definitions of treatment response do not account for patient compliance, which biomarkers cannot reliably distinguish from physiological drug resistance. - Clinicians questioned the practical utility of such a tool, noting that it remains unclear what specific intervention or action a biomarker result would trigger. | N/A |  | Keep in the final framework |
| Test-of-cure (93%) | - Comments were largely related to the difficulty of test-of-cure studies providing definitive answers, and the lack of validated biomarkers currently available. | N/A |  | Keep in the final framework |
| Post-TB lung disease (83%) | - Distinguishing between relapse and reinfection does not alter immediate clinical management, as the individual is treated the same way regardless of the cause. - Clinicians prioritize determining the drug resistance profile and appropriate treatment duration over identifying the source of the recurrence. - The value of differentiating relapse from reinfection is primarily academic, serving research, public health surveillance, and epidemiology rather than routine care. - Current methods, such as Whole Genome Sequencing (WGS), are considered sufficient for this purpose, making a new dedicated biomarker a low priority. | N/A |  | Keep in the final framework |
| Relapse or re-infection (79%) | - Distinguishing between relapse and reinfection does not alter immediate clinical management, as the individual is treated the same way regardless of the cause. - Clinicians prioritize determining the drug resistance profile and appropriate treatment duration over identifying the source of the recurrence. - The value of differentiating relapse from reinfection is primarily academic, serving research, public health surveillance, and epidemiology rather than routine care. - Current methods, such as Whole Genome Sequencing (WGS), are considered sufficient for this purpose, making a new dedicated biomarker a low priority. | - Whole Genome Sequencing (WGS) already effectively differentiates between the two by comparing bacterial strains from separate episodes. - The distinction has minimal impact on immediate patient care, as the priority remains treating the recurrence and addressing drug resistance. - In resource-constrained settings, this use case is a lower priority compared to more urgent needs like treatment response or viable infection biomarkers. - While less critical for patient management, the distinction remains important for tracking epidemiological trends and evaluating population-level treatment efficacy. | Rejected through the Mentimeter poll, with 27 to 9 voting to omit the use case from the final framework. | Omit from the final framework |

### S11 Table: The percentage of interest-holders in each group who agreed with the use cases.

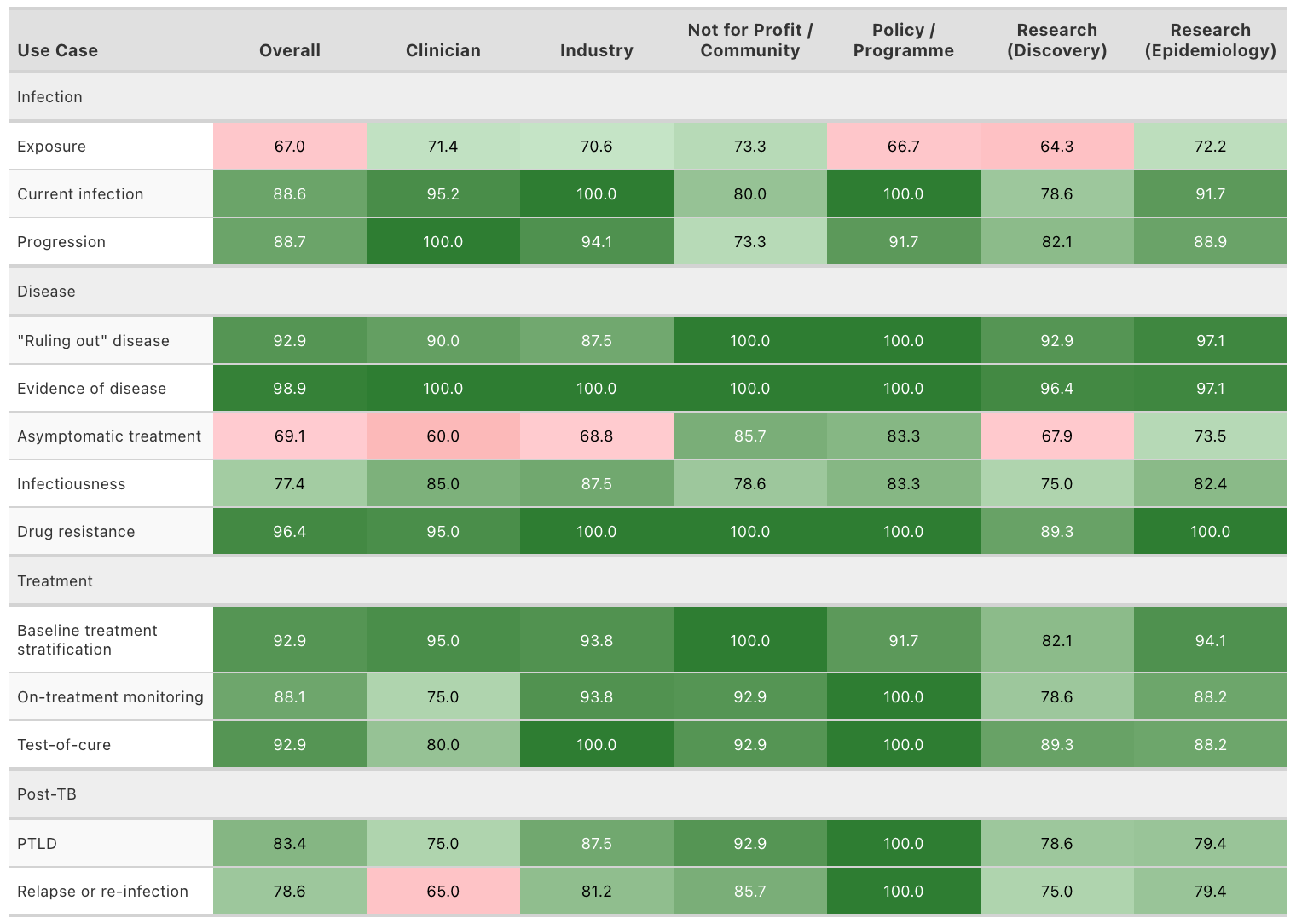

Values represent the percentage of respondents who selected “Somewhat Agree” or “Strongly Agree” for each use case. Color coding indicates the level of consensus, with 70% being the threshold: green cells (≥70%) represent strong agreement, while red cells (<70%) indicate less consensus. Percentages are calculated based on the total number of times that interest-holder group was selected, not the total number of respondents.
S12 Table: The importance of use cases for TB biomarkers and tests by interest-holder groups.

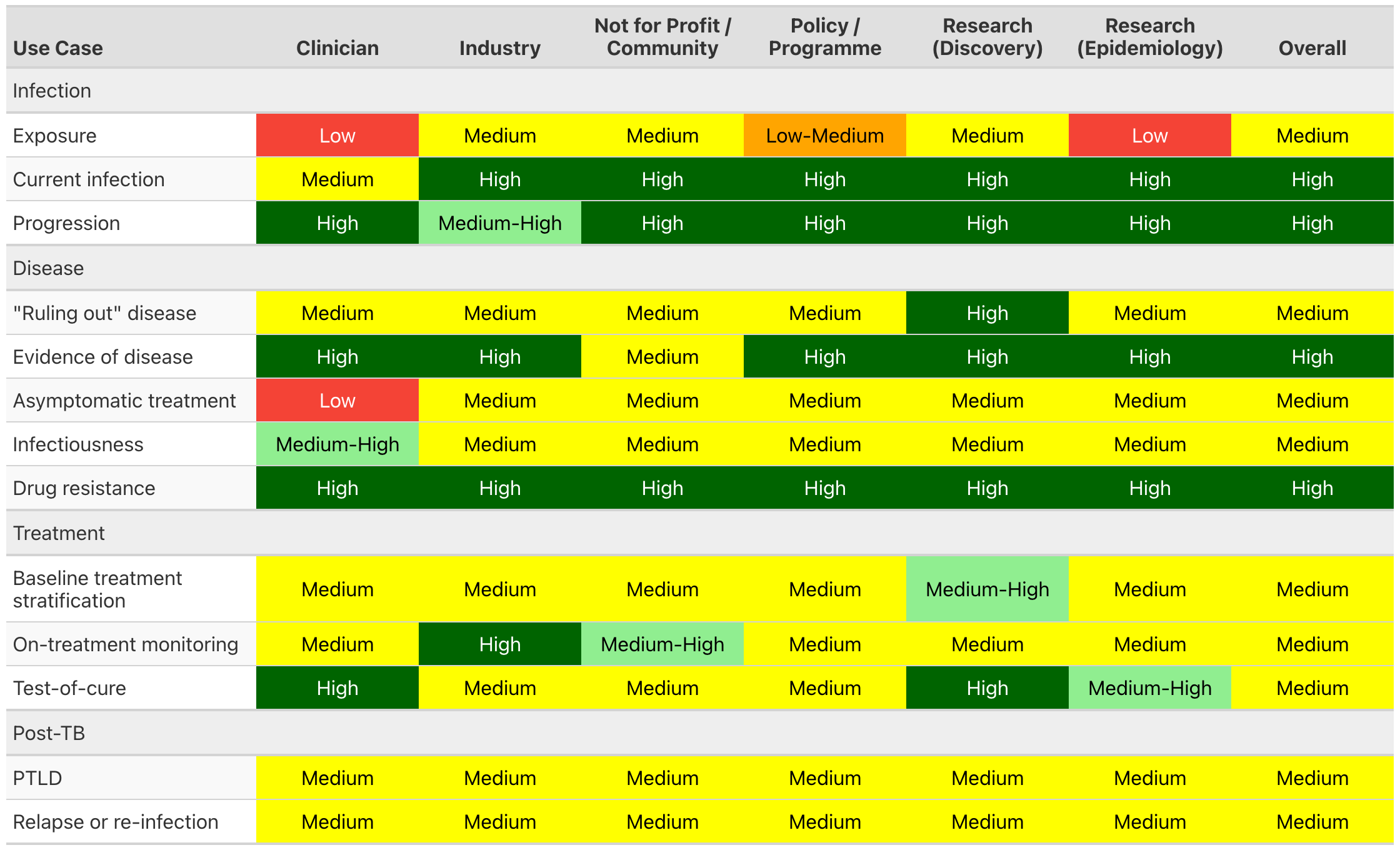

For each interest-holder group, the reported importance value is the mode of responses within that group. When responses are tied (equal number of respondents selecting different importance levels), they are combined (e.g., “Low-Medium”). Specific cases like “Low-High” and “Low-Medium-High” are converted to “Medium”. The “Overall” column represents the most commonly selected importance level across all individual respondents, regardless of interest-holder group.

PRISMA-ScR Checklist

| **SECTION** | **ITEM** | **PRISMA-ScR CHECKLIST ITEM** | **REPORTED ON PAGE #** |
| --- | --- | --- | --- |
| **TITLE** | | | |
| Title | 1 | Identify the report as a scoping review. | N/A |
| **ABSTRACT** | | | |
| Structured summary | 2 | Provide a structured summary that includes (as applicable): background, objectives, eligibility criteria, sources of evidence, charting methods, results, and conclusions that relate to the review questions and objectives. | Protocol (p.1, “Abstract”) |
| **INTRODUCTION** | | | |
| Rationale | 3 | Describe the rationale for the review in the context of what is already known. Explain why the review questions/objectives lend themselves to a scoping review approach. | Introduction (p.2)  Protocol (p.2) |
| Objectives | 4 | Provide an explicit statement of the questions and objectives being addressed with reference to their key elements (e.g., population or participants, concepts, and context) or other relevant key elements used to conceptualize the review questions and/or objectives. | Methods (p.2, “Scoping review”)  Protocol (p.2, “Introduction”) |
| **METHODS** | | | |
| Protocol and registration | 5 | Indicate whether a review protocol exists; state if and where it can be accessed (e.g., a Web address); and if available, provide registration information, including the registration number. | Methods (p.2) |
| Eligibility criteria | 6 | Specify characteristics of the sources of evidence used as eligibility criteria (e.g., years considered, language, and publication status), and provide a rationale. | Supplementary table 1 |
| Information sources* | 7 | Describe all information sources in the search (e.g., databases with dates of coverage and contact with authors to identify additional sources), as well as the date the most recent search was executed. | Supplementary table table 1 |
| Search | 8 | Present the full electronic search strategy for at least 1 database, including any limits used, such that it could be repeated. | Supplementary table 1 |
| Selection of sources of evidence† | 9 | State the process for selecting sources of evidence (i.e., screening and eligibility) included in the scoping review. | Methods (p. 2, “Scoping review”) |
| Data charting process‡ | 10 | Describe the methods of charting data from the included sources of evidence (e.g., calibrated forms or forms that have been tested by the team before their use, and whether data charting was done independently or in duplicate) and any processes for obtaining and confirming data from investigators. | Methods (p.2, “Scoping review”)  Protocol (p.3, “Data extraction and analysis”) |
| Data items | 11 | List and define all variables for which data were sought and any assumptions and simplifications made. | Supplementary table 4 |
| Critical appraisal of individual sources of evidence | 12 | If done, provide a rationale for conducting a critical appraisal of included sources of evidence; describe the methods used and how this information was used in any data synthesis (if appropriate). | N/A |
| Synthesis of results | 13 | Describe the methods of handling and summarizing the data that were charted. | Methods (p. 2, “Scoping review”)  Protocol (p. 3, “Data extraction and analysis”) |
| **RESULTS** | | | |
| Selection of sources of evidence | 14 | Give numbers of sources of evidence screened, assessed for eligibility, and included in the review, with reasons for exclusions at each stage, ideally using a flow diagram. | Results (p. 4, “Scoping review”)  Supplementary figure 1 |
| Characteristics of sources of evidence | 15 | For each source of evidence, present characteristics for which data were charted and provide the citations. | N/A |
| Critical appraisal within sources of evidence | 16 | If done, present data on critical appraisal of included sources of evidence (see item 12). | N/A |
| Results of individual sources of evidence | 17 | For each included source of evidence, present the relevant data that were charted that relate to the review questions and objectives. | Supplementary table 7 |
| Synthesis of results | 18 | Summarize and/or present the charting results as they relate to the review questions and objectives. | Results (p.4, 5, 6, 7 “Use cases for TB biomarkers and tests by TB care pathway stage”)  Table 1  Supplementary table 7, |
| **DISCUSSION** | | | |
| Summary of evidence | 19 | Summarize the main results (including an overview of concepts, themes, and types of evidence available), link to the review questions and objectives, and consider the relevance to key groups. | Discussion (p. 8) |
| Limitations | 20 | Discuss the limitations of the scoping review process. | Discussion (p.9) |
| Conclusions | 21 | Provide a general interpretation of the results with respect to the review questions and objectives, as well as potential implications and/or next steps. | Discussion (p.9) |
| **FUNDING** | | | |
| Funding | 22 | Describe sources of funding for the included sources of evidence, as well as sources of funding for the scoping review. Describe the role of the funders of the scoping review. | N/A |

JBI = Joanna Briggs Institute; PRISMA-ScR = Preferred Reporting Items for Systematic reviews and Meta-Analyses extension for Scoping Reviews.

* Where *sources of evidence* (see second footnote) are compiled from, such as bibliographic databases, social media platforms, and Web sites.

† A more inclusive/heterogeneous term used to account for the different types of evidence or data sources (e.g., quantitative and/or qualitative research, expert opinion, and policy documents) that may be eligible in a scoping review as opposed to only studies. This is not to be confused with *information sources* (see first footnote).

‡ The frameworks by Arksey and O’Malley (6) and Levac and colleagues (7) and the JBI guidance (4, 5) refer to the process of data extraction in a scoping review as data charting*.*

§ The process of systematically examining research evidence to assess its validity, results, and relevance before using it to inform a decision. This term is used for items 12 and 19 instead of "risk of bias" (which is more applicable to systematic reviews of interventions) to include and acknowledge the various sources of evidence that may be used in a scoping review (e.g., quantitative and/or qualitative research, expert opinion, and policy document).

*From:* Tricco AC, Lillie E, Zarin W, O'Brien KK, Colquhoun H, Levac D, et al. PRISMA Extension for Scoping Reviews (PRISMAScR): Checklist and Explanation. Ann Intern Med. 2018;169:467–473. [doi: 10.7326/M18-0850](http://annals.org/aim/fullarticle/2700389/prisma-extension-scoping-reviews-prisma-scr-checklist-explanation).

### ACCORD (ACcurate COnsensus Reporting Document) Checklist

| Item No. | Section | Checklist Item (*help text*) | Page No. |
| --- | --- | --- | --- |
| T1 | **Title** | Identify the article as reporting a consensus exercise and state the consensus methods used in the title.  *For example, Delphi or nominal group technique.* | Title |
| I1 | **Introduction** | Explain why a consensus exercise was chosen over other approaches. | Introduction (P. 1) |
| I2 |  | State the aim of the consensus exercise, including its intended audience and geographical scope (national, regional, global). | Introduction (P. 1) |
| I3 |  | If the consensus exercise is an update of an existing document, state why an update is needed, and provide the citation for the original document. | N/A |
| M1 | **Methods**  Registration | If the study or study protocol was prospectively registered, state the registration platform and provide a link. If the exercise was not registered, this should be stated.  *Recommended to include the date of registration.* | Methods (P.2) |
| M2 | Selection of SC and/or panellists | Describe the role(s) and areas of expertise or experience of those directing the consensus exercise.  *For example, whether the project was led by a chair, co-chairs or a steering committee, and, if so, how they were chosen. List their names if appropriate, and whether there were any subgroups for individual steps in the process.* | Methods (P.2 “Participants/interest-holders”)  Supplementary table 5 |
| M3 |  | Explain the criteria for panellist inclusion and the rationale for panellist numbers. State who was responsible for panellist selection. | Methods (P.2 “Participants/interest-holders”) |
| M4 |  | Describe the recruitment process (how panellists were invited to participate).  *Include communication/advertisement method(s) and locations, numbers of invitations sent, and whether there was centralised oversight of invitations or if panellists were asked/allowed to suggest other members of the panel.* | Methods (P.2 “Participants/interest-holders”)  Figure 1 |
| M5 |  | Describe the role of any members of the public, patients or carers in the different steps of the study. | N/A |
| M6 | Preparatory research | Describe how information was obtained prior to generating items or other materials used during the consensus exercise.  *This might include a literature review, interviews, surveys, or another process.* | Methods (P.2 “Scoping review”) |
| M7 |  | Describe any systematic literature search in detail, including the search strategy and dates of search or the citation if published already.  *Provide the details suggested by the reporting guideline PRISMA and the related PRISMA-Search extension.* | Methods (p.2 “Scoping review”)  Supplementary tables 1, 2, 3, 4 Protocol (p.2 and p.3, “Scoping review”) |
| M8 |  | Describe how any existing scientific evidence was summarised and if this evidence was provided to the panellists. | Methods (p.2 “Scoping review”)  Methods (p.3 “Consensus process”) |
| M9 | Assessing consensus | Describe the methods used and steps taken to gather panellist input and reach consensus (for example, Delphi, RAND-UCLA, nominal group technique).  *If modifications were made to the method in its original form, provide a detailed explanation of how the method was adjusted and why this was necessary for the purpose of your consensus-based study.* | Methods (p.3 “Consensus process”)  Figure 1 |
| M10 |  | Describe how each question or statement was presented and the response options. State whether panellists were able to or required to explain their responses, and whether they could propose new items.  *Where possible, present the questionnaire or list of statements as supplementary material.* | Supplementary Table 6 |
| M11 |  | State the objective of each consensus step.  *A step could be a consensus meeting, a discussion or interview session, or a Delphi round.* | Methods (p.3, “Consensus process”) Figure 1 |
| M12 |  | State the definition of consensus (for example, number, percentage, or categorical rating, such as ‘agree’ or ‘strongly agree’) and explain the rationale for that definition. | Methods (p.3, “Consensus process”) |
| M13 |  | State whether items that met the prespecified definition of consensus were included in any subsequent voting rounds. | Methods (p.3, “Consensus process”) |
| M14 |  | For each step, describe how responses were collected, and whether responses were collected in a group setting or individually. | Methods (p.3, “Consensus process”) |
| M15 |  | Describe how responses were processed and/or synthesised.  *Include qualitative analyses of free-text responses (for example, thematic, content or cluster analysis) and/or quantitative analytical methods, if used.* | Methods (p.3, “Consensus process”) |
| M16 |  | Describe any piloting of the study materials and/or survey instruments.  *Include how many individuals piloted the study materials, the rationale for the selection of those individuals, any changes made as a result and whether their responses were used in the calculation of the final consensus. If no pilot was conducted, this should be stated.* | N/A |
| M17 |  | If applicable, describe how feedback was provided to panellists at the end of each consensus step or meeting.  *State whether feedback was quantitative (for example, approval rates per topic/item) and/or qualitative (for example, comments, or lists of approved items), and whether it was anonymised.* | N/A |
| M18 |  | State whether anonymity was planned in the study design. Explain where and to whom it was applied and what methods were used to guarantee anonymity. | Methods (p.3, “Consensus process”) |
| M19 |  | State if the steering committee was involved in the decisions made by the consensus panel.  *For example, whether the steering committee or those managing consensus also had voting rights.* | Methods (p.3, “Consensus process”) |
| M20 | Participation | Describe any incentives used to encourage responses or participation in the consensus process.  *For example, were invitations to participate reiterated, or were participants reimbursed for their time.* | Methods (p.3) |
| M21 |  | Describe any adaptations to make the surveys/meetings more accessible.  *For example, the languages in which the surveys/meetings were conducted and whether translations or plain language summaries were available*. | N/A |
| R1 | Results | State when the consensus exercise was conducted. List the date of initiation and the time taken to complete each consensus step, analysis, and any extensions or delays in the analysis. | Methods (p.3, “Consensus process”)  Results (p.4 “Consensus process”) |
| R2 |  | Explain any deviations from the study protocol, and why these were necessary.  *For example, addition of panel members during the exercise, number of consensus steps, stopping criteria; report the step(s) in which this occurred.* | Methods (p.3, “Consensus process”) |
| R3 |  | For each step, report quantitative (number of panellists, response rate) and qualitative (relevant socio-demographics) data to describe the participating panellists. | Figure 1  Results (p.4 “Consensus process”)  Supplementary table 9 |
| R4 |  | Report the final outcome of the consensus process as qualitative (for example, aggregated themes from comments) and/or quantitative (for example, summary statistics, score means, medians and/or ranges) data. | Figure 2  Table 2  Supplementary table 8  Supplementary table 10 |
| R5 |  | List any items or topics that were modified or removed during the consensus process. Include why and when in the process they were modified or removed. | N/A |
| D1 | Discussion | Discuss the methodological strengths and limitations of the consensus exercise.  *Include factors that may have impacted the decisions (for example, response rates, representativeness of the panel, potential for feedback during consensus to bias responses, potential impact of any non-anonymised interactions).* | Discussion (p.9) |
| D2 |  | Discuss whether the recommendations are consistent with any pre-existing literature and, if not, propose reasons why this process may have arrived at alternative conclusions. | Discussion (p.8) |
| O1 | Other information | List any endorsing organisations involved and their role. | See funding statement. No other endorsing organisations |
| O2 |  | State any potential conflicts of interests, including among those directing the consensus study and panellists. Describe how conflicts of interest were managed. | Methods P3, See also protocol |
| O3 |  | State any funding received and the role of the funder.  *Specify, for example, any funder involvement in the study concept/design, participation in the steering committee, conducting the consensus process, funding of any medical writing support. This could be disclosed in the methods or in the relevant transparency section of the manuscript. Where a funder did not play a role in the process or influence the decisions reached, this should be specified.* | See funding statement |

For more information see: <https://www.ismpp.org/accord> and Gattrell WT, Logullo P, van Zuuren EJ, Price A, Hughes EL, Blazey P, et al. (2024) ACCORD (ACcurate COnsensus Reporting Document): A reporting guideline for consensus methods in biomedicine developed via a modified Delphi. PLoS Med 21(1): e1004326. https://doi.org/10.1371/journal.pmed.1004326
